# Development and testing of a polygenic risk score for breast cancer aggressiveness

**DOI:** 10.1101/2022.08.26.22278957

**Authors:** Yiwey Shieh, Jacquelyn Roger, Christina Yau, Denise M. Wolf, Gillian L. Hirst, Lamorna Brown Swigart, Scott Huntsman, Donglei Hu, Jovia L. Nierenberg, Pooja Middha, Rachel S. Heise, Yushu Shi, Linda Kachuri, Qianqian Zhu, Song Yao, Christine B. Ambrosone, Marilyn L. Kwan, Bette J. Caan, John S. Witte, Lawrence H. Kushi, Laura van ‘T Veer, Laura J. Esserman, Elad Ziv

## Abstract

Aggressive breast cancers portend a poor prognosis, but current polygenic risk scores (PRSs) for breast cancer do not reliably predict aggressive cancers. Aggressiveness can be effectively recapitulated using tumor gene expression profiling. Thus, we sought to develop a novel PRS for the risk of recurrence score weighted on proliferation (ROR-P), an established prognostic signature. Using 2,363 breast cancers with tumor gene expression data and single nucleotide polymorphism (SNP) genotypes, we examined the associations between ROR-P and known breast cancer susceptibility SNPs using linear regression models. We constructed PRSs based on varying p-value thresholds and selected the optimal PRS based on model r^2^ in 10-fold cross-validation. We then used Cox proportional hazards regression to test the ROR-P PRS’s association with breast cancer-specific survival in two independent cohorts totaling 10,196 breast cancers and 785 events. In meta-analysis of these cohorts, higher ROR-P PRS was associated with worse survival, HR per SD = 1.13 (95% CI 1.05-1.21, p=0.001). The ROR-P PRS had a similar magnitude of effect on survival as a comparator PRS for estrogen receptor (ER)-negative versus positive cancer risk (PRS_ER-/ER+)._ Furthermore, its effect was minimally attenuated when adjusted for PRS_ER-/ER+_, suggesting that the ROR-P PRS provides additional prognostic information beyond ER status. In summary, we used a novel approach based on integrated analysis of germline SNP and tumor gene expression data to construct a PRS associated with aggressive tumor biology and worse survival. These findings could potentially enhance risk stratification for breast cancer screening and prevention.

## Introduction

Polygenic risk scores (PRSs) have emerged as promising tools for breast cancer risk prediction. Over 200 single nucleotide polymorphisms (SNPs) associated with breast cancer risk have been identified^1^. Though the effects of individual SNPs are weak, PRSs representing the cumulative effects of multiple SNPs can stratify breast cancer risk on a population level^2^ and improve the performance of clinical breast cancer risk prediction models^3,4^. Ongoing prospective trials are testing the ability of the PRS to inform decision-making around breast cancer screening and prevention^5-7^.

Current breast cancer PRSs have limited ability to account for the biological heterogeneity of breast cancer. This is a critical limitation since breast cancer encompasses a variety of subtypes ranging from indolent to aggressive, with the latter defined as having increased proliferation or metastatic potential and poor prognosis^8^. However, case-only analyses have found that PRSs for overall breast cancer risk are associated with more favorable clinicopathologic^9^ and prognostic features^10^, as well as lower risk of interval versus screen-detected cancer^11,12^. Efforts to fit PRSs to subtypes of breast cancer have focused on estrogen receptor (ER) status given that ER-negative breast cancers tend to be more proliferative and are associated with earlier risk of relapse^2,13^. However, aggressive cancers can encompass ER-negative and ER-positive subtypes. For instance, ER-positive cancers are commonly divided into luminal A (low-grade) and B (high-grade) subtypes with the latter having worse prognosis^14^.

Beyond ER status, aggressiveness can be measured using tumor prognostic signatures, which integrate the expression levels of multiple genes to calculate prognostic scores that guide treatment decision-making^8,15,16^. In this analysis, we selected the risk of recurrence score weighted on proliferation (ROR-P), which is based on the expression of 50 genes included in the Prediction Analysis of Microarray 50 (PAM50) signature. PAM50 classifies tumors by intrinsic subtype (luminal A, luminal B, HER2-enriched, basal-like, and normal-like). ROR-P is calculated by adding the subtype-centroid correlation coefficients for each subtype, weighted by their association with recurrence, to the weighted expression levels of 11 proliferation genes^17,18^. ROR-P is continuous (though categorical cutoffs are used for clinical decision-making) and has stronger prognostic value than traditional markers such as ER status, grade, and Ki-67^17,19^.

Prior studies have examined the associations between germline genetics and ROR-P using a transcriptome-wide association study approach^20^. However, no studies have attempted to develop a PRS for ROR-P or other gene expression-based signature of aggressiveness. We hypothesized that some known breast cancer susceptibility SNPs are positively correlated with ROR-P, whereas others are negatively correlated, and these differential associations can be used to construct a PRS for ROR-P. We therefore sought to construct the ROR-P PRS in datasets with germline SNP genotypes and tumor gene expression. To evaluate whether the ROR-P PRS was associated with aggressive tumors with worse prognosis, we examined its association with breast cancer-specific survival in two independent cohorts.

## Results

### Study characteristics

We developed the ROR-P PRS using 2,363 breast cancers from three studies: The Cancer Genome Atlas (TCGA)^21^, Molecular Taxonomy of Breast Cancer International Consortium (METABRIC)^22^, and Investigation of Serial Studies to Predict Your Therapeutic Response with Imaging And molecular analysis 2 (I-SPY 2 TRIAL)^23^ (**Figure 1, Table S1, Figure S1**). Most of the participants in these studies were non-Hispanic white; METABRIC did not report race or ethnicity but predominantly included white participants because recruitment occurred in the United Kingdom and Canada^22^ (**Table 1**). Cancers from I-SPY 2 were diagnosed at younger ages and more likely to be ROR-P High and classified as Basal intrinsic subtype. This reflects the trial’s inclusion criteria, which is limited to locally advanced, molecularly high-risk cancers as defined by gene expression profiling and/or clinicopathologic features^23^.

**Table 1.**
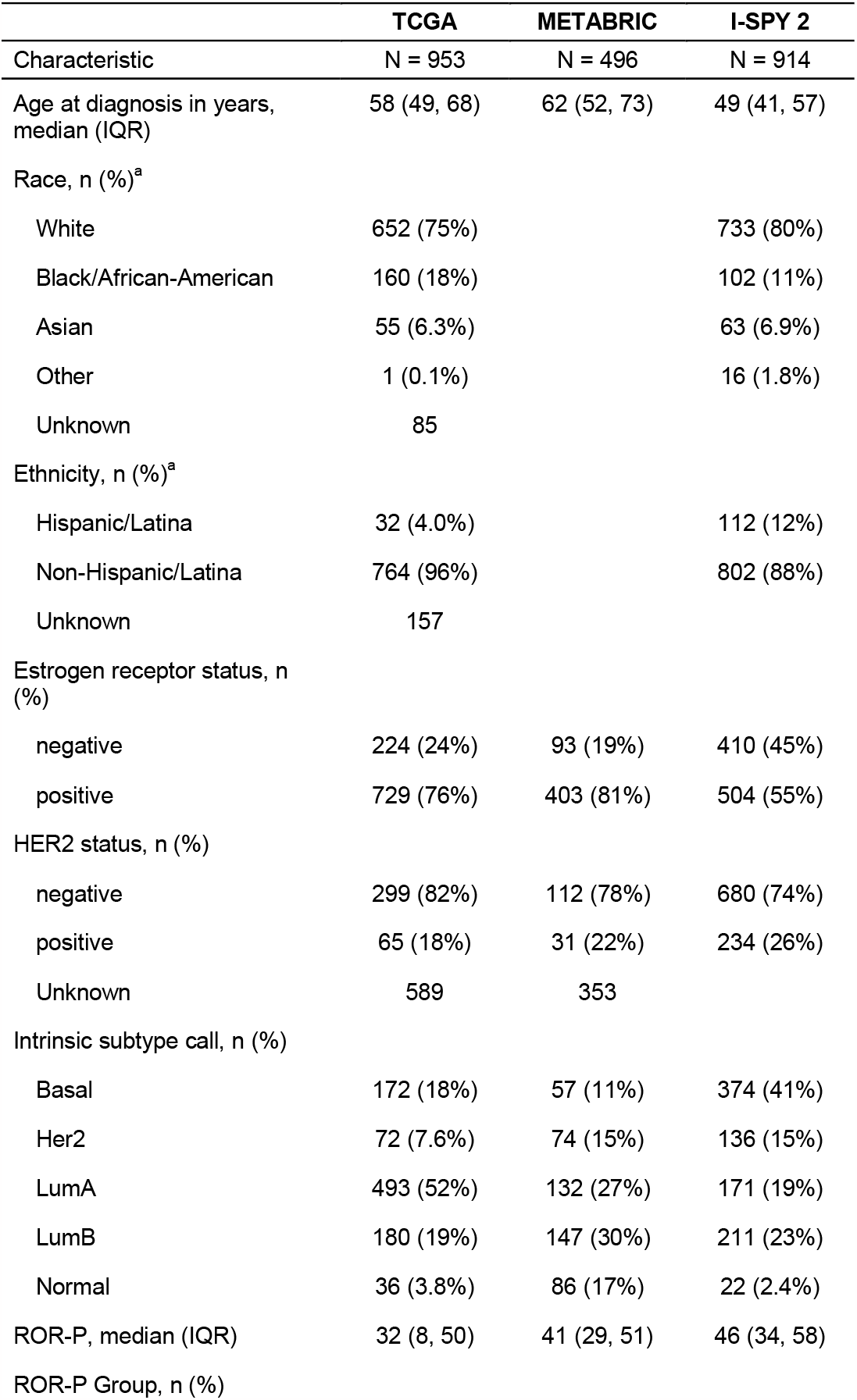
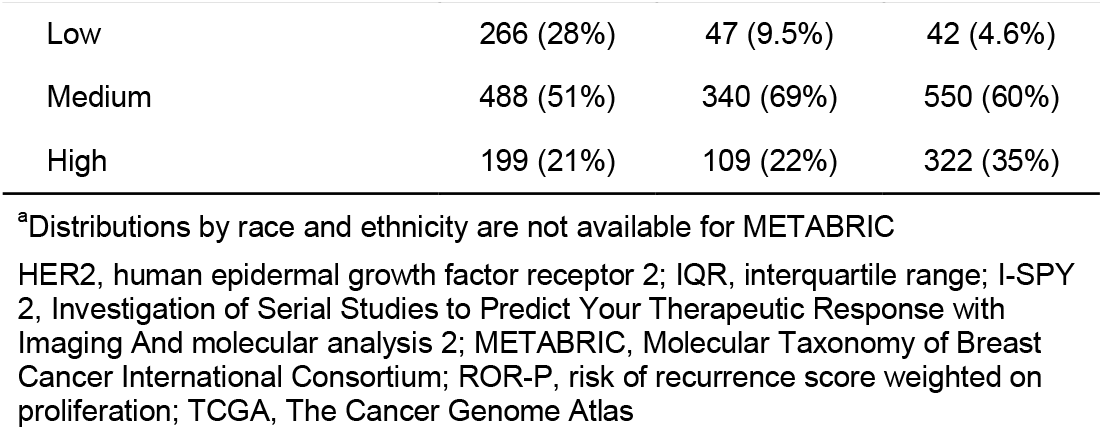
Characteristics of studies used in development of ROR-P PRS

**Figure 1.**
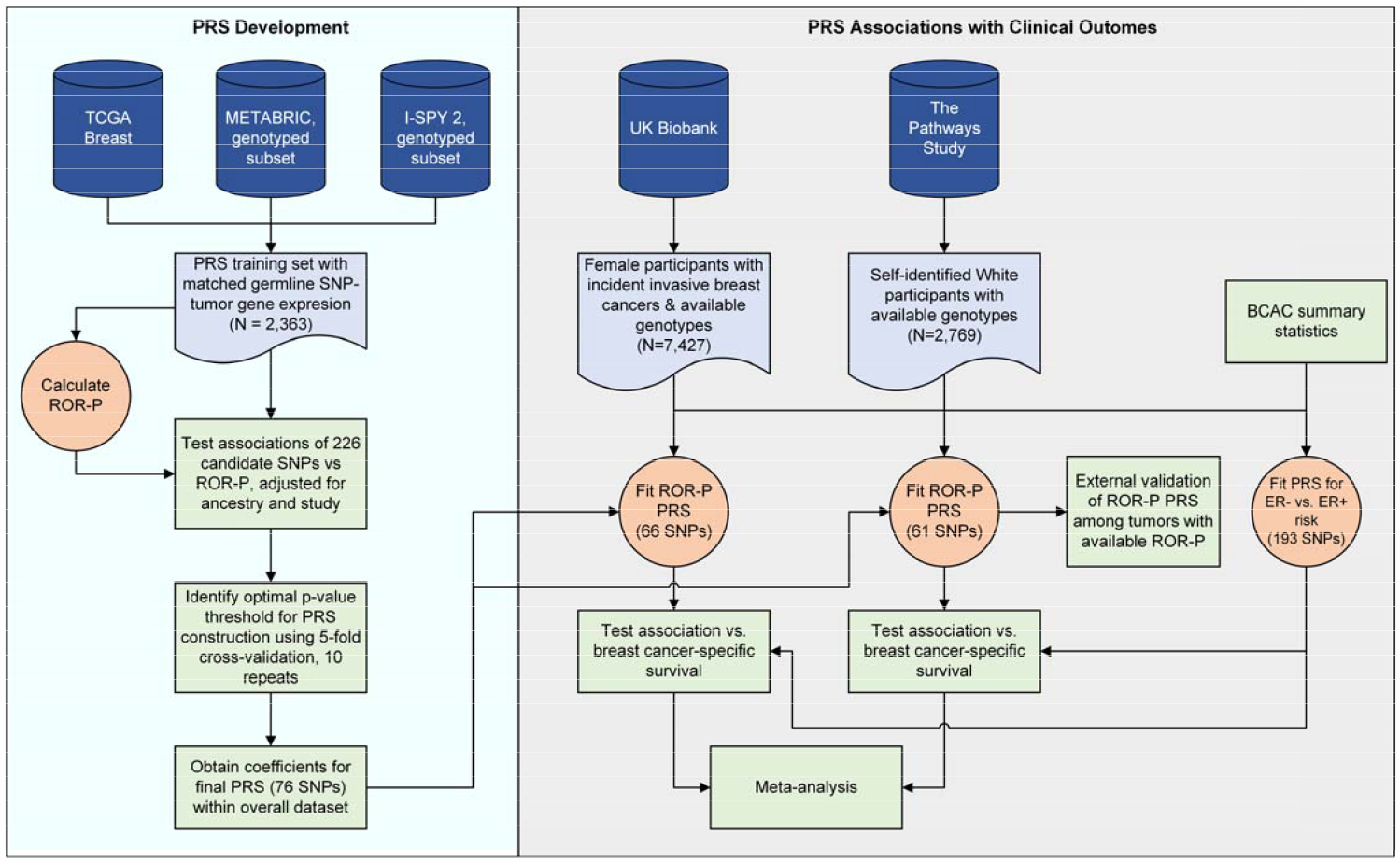
Study design. We developed a polygenic risk score (PRS) for the risk of recurrence score weighted on proliferation (ROR-P) using pooled data from three studies: The Cancer Genome Atlas (TCGA), Molecular Taxonomy of Breast Cancer International Consortium (METABRIC), and Investigation of Serial Studies to Predict Your Therapeutic Response with Imaging And molecular analysis 2 (I-SPY 2 TRIAL). We used these datasets to evaluate the performance of PRS constructed using varying p-value thresholds and to calculate the effect sizes of the SNPs included in the PRS with optimal performance (cross-validated r^2^). We then calculated the ROR-P PRS in breast cancer patients from two independent datasets, the UK Biobank and the Pathways Study. Within Pathways, we performed external validation of the ROR-P PRS by examining its association with measured ROR-P within tumors with available gene expression profiling data. We then tested the associations between the ROR-P PRS and breast cancer-specific survival in UK Biobank and the Pathways Study and performed meta-analysis of the results. In parallel, we generated a PRS for the case-case risk of estrogen receptor (ER)-negative versus ER-positive breast cancer using summary statistics from the Breast Cancer Association Consortium (BCAC) and evaluated its association with breast cancer-specific survival.

We calculated the ROR-P PRS and tested its associations with survival and tumor characteristics in two studies containing prospective follow-up of breast cancer patients, the UK Biobank and the Pathways Study (**Figure 1, Table S2**). Limited breast cancer characteristics were available for UK Biobank, but age at diagnosis and event rate were comparable across studies, with the Pathways Study having a longer duration of follow-up (**Table S3**). Our analysis of UK Biobank included 7, 427 breast cancer cases and 544 breast cancer-specific deaths during a median follow-up time of 6.4 (interquartile range 3.7-9.1) years. The Pathways Study included 2,769 cancers with 241 breast cancer-specific deaths during a median follow-up time of 10.7 (interquartile range 8.2-12.3) years. Tumors in Pathways were predominantly ER-positive, human epidermal growth factor receptor 2 (HER2)-negative, and Grade I or II.

### Development of the ROR-P PRS

Using pooled data from TCGA, METABRIC, and the I-SPY 2 TRIAL, we evaluated the case-case associations of 226 breast cancer susceptibility SNPs and ROR-P (**Table S4**). Based on these associations, we constructed PRSs with varying p-value thresholds and identified a 76-SNP PRS as having the best performance, with a model r^2^ of 0.051 in 5-fold cross-validation (**Figure 2A, Table S5**). For 51 of 76 SNPs in the ROR-P PRS, the breast cancer risk allele, as annotated by the original GWAS, was associated with lower ROR-P (**Table S6, Figure 2B**), which is consistent with the observation that the PRS for overall breast cancer risk is associated with better survival^10^. Six of 76 SNPs in our final ROR-P PRS had nominally significant associations with ROR-P, though none met significance after Bonferroni correction. The SNP with the strongest positive association with ROR-P, rs67397200 (*ABHD8* in 19p13.11), was discovered in GWAS for ER-negative cancer^13^ and is associated with increased case-case risk of luminal B and triple negative intrinsic-like subtypes, and decreased risk of luminal A subtype^24^. The SNP with the strongest negative association, rs537626 (*LINC01488* in 11q13.3), was discovered in GWAS for early onset breast cancer^25^, although the region has also been implicated in increased risk of ER-positive cancer in an admixture mapping study of African-American women.^26^

**Figure 2.**
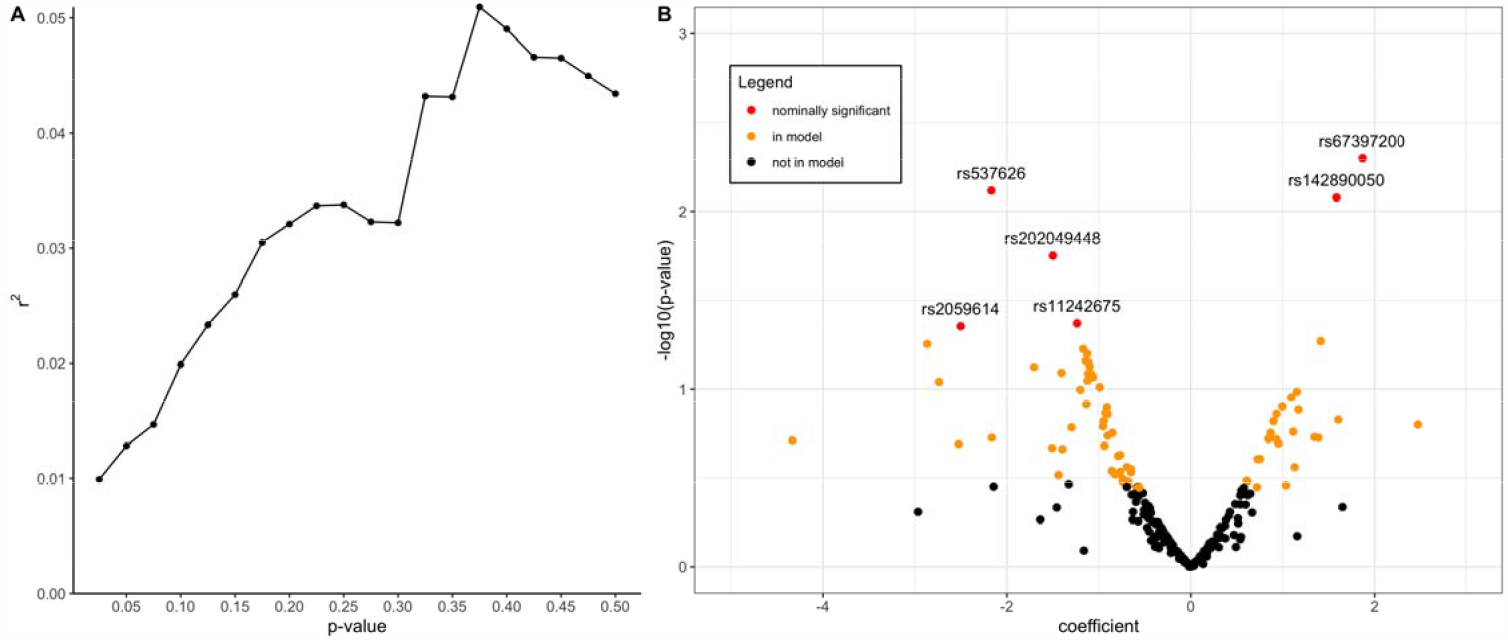
Development of the polygenic risk score for the risk of recurrence score weighted on proliferation (ROR-P PRS) A. We performed 5-fold cross-validation to identify the optimal p-value threshold for including single nucleotide polymorphisms (SNPs) in the ROR-P PRS. The r^2^ is shown for each p-value threshold tested. Our final PRS used a p-value cutoff of 0.375. B. Volcano plot of associations of 226 SNPs with ROR-P, adjusted for genetic ancestry (principal components 1-10) and study. SNPs included in the final 76-SNP PRS score are indicated in red (nominally significant association with ROR-P) and orange (not nominally significant but included in model).

### External validation of ROR-P PRS

We performed external validation of the ROR-P PRS in a nested subset of 484 tumors from the Pathways Study that had undergone gene expression profiling^27^ (**Figure S2**). The ROR-P PRS was weakly correlated with measured ROR-P, with a Pearson correlation coefficient of 0.095 (p=0.037) (**Figure 3A**). We also examined associations between the ROR-P PRS and tumor clinicopathologic features (**Figure 3B**). Higher ROR-P PRS was associated with ER-negative status, odds ratio per standard deviation increment (OR per S.D.) of 1.13 (95% CI 1.01-1.26), but not HER2 status or grade. Higher ROR-P PRS was also associated with increased odds of basal versus luminal A intrinsic-like subtype, as defined by receptor status and grade (OR per S.D. = 1.19, 95% CI 1.05-1.35, p=0.007).

**Figure 3.**
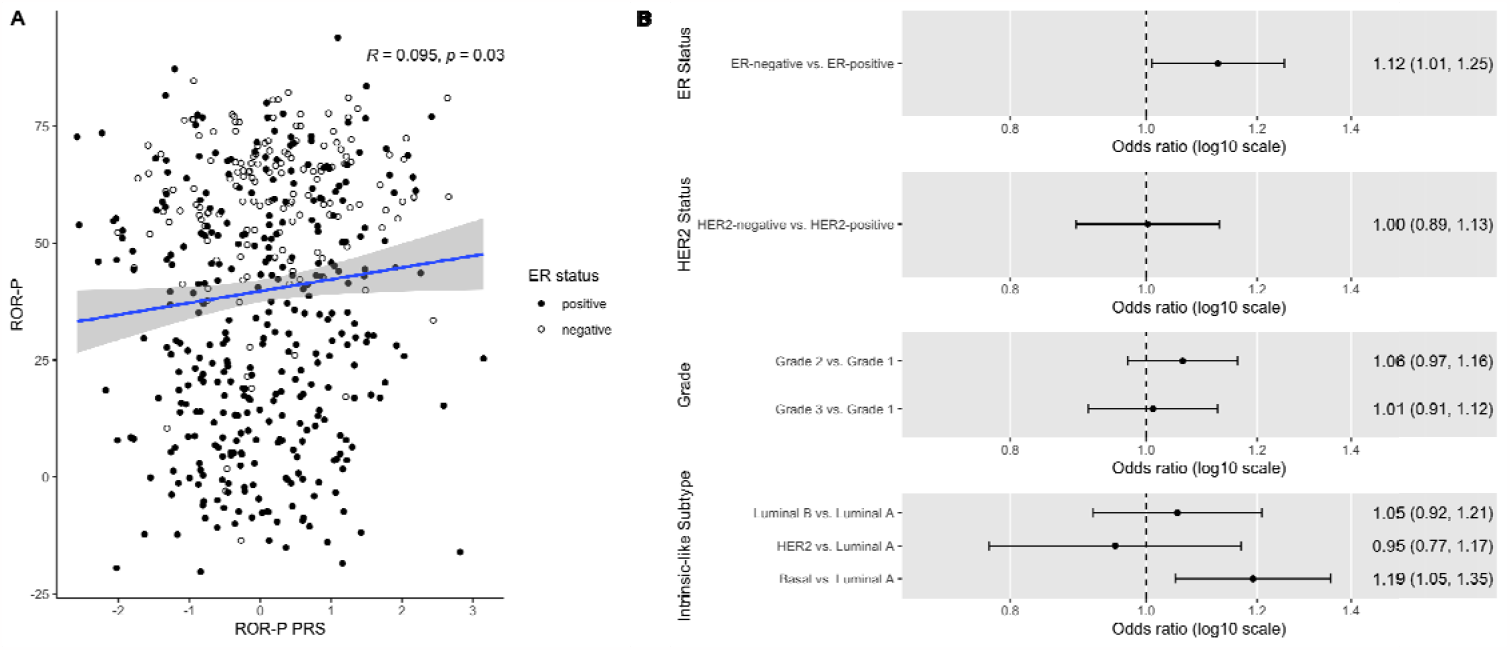
Validation of a polygenic risk score for risk of recurrence score weighted on proliferation (ROR-P PRS) in the Pathways Study. A. Scatter plot of the normalized ROR-P PRS and tumor ROR-P in a subset of 484 tumors with both available measures. The Pearson correlation coefficient and p-value are shown. Points are color-coded by estrogen receptor (ER) status. B. The associations between ROR-P PRS and tumor features in the Pathways Study were evaluated using logistic regression (for estrogen receptor and human epidermal growth factor 2 status) and multinomial logistic regression (for histologic grade and intrinsic-like subtype). Models were adjusted for genetic ancestry (principal components 1-10). Point estimates and 95% confidence intervals for the association between ROR-P PRS and respective tumor feature are shown.

### Association of ROR-P PRS with breast cancer-specific survival

In UK Biobank, we first evaluated the association between a PRS for overall breast cancer risk (PRS_overallBC_) and breast cancer-specific survival. We hypothesized that since prior studies have shown that the overall breast cancer PRS is associated with favorable characteristics in breast cancer patients^10^, then higher overall breast cancer PRS among cases would be associated with more favorable survival. As expected, PRS_overallBC_ was inversely associated with breast cancer mortality, with a hazard ratio per standard deviation (HR per S.D.) of 0.86 (95% CI 0.78-0.94, p=0.019). In Kaplan-Meier analyses, the bottom tertile of the PRS, corresponding with the lowest risk of developing breast cancer, was associated with worst survival (**Figure 4**).

**Figure 4.**
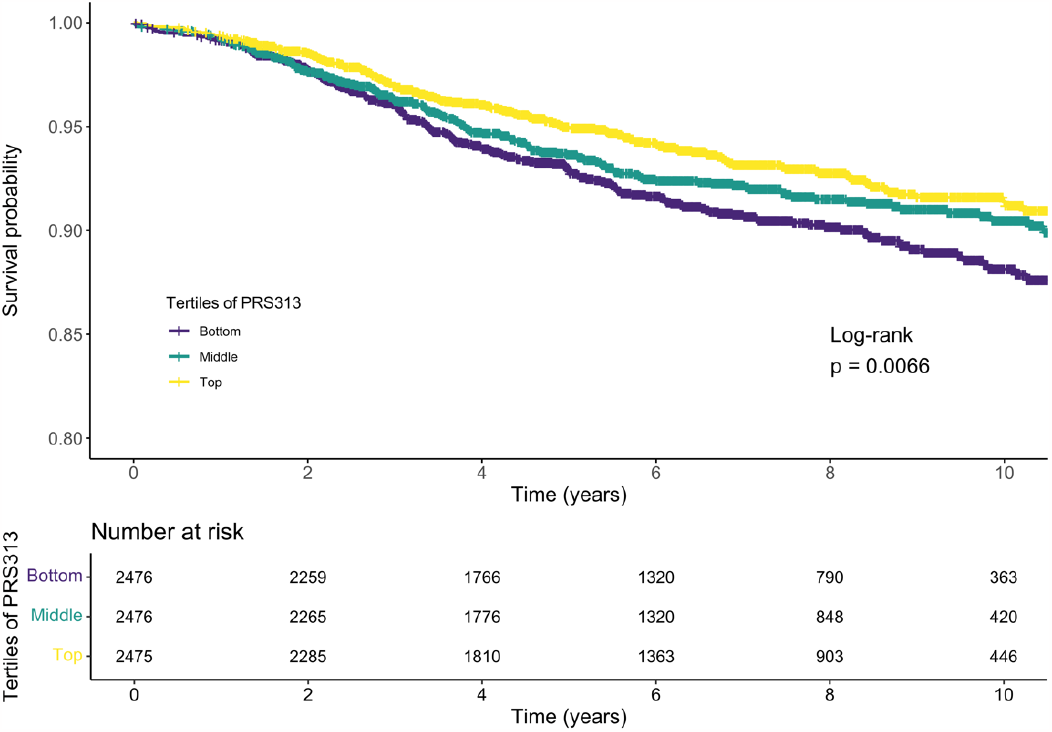
Association between a polygenic risk score (PRS_overallBC_) for overall invasive breast cancer risk and breast cancer-specific survival. Kaplan-Meier survival analysis of tertiles of a 274-SNP PRS for overall breast cancer risk in the UK Biobank. p-values for the log-rank test are shown.

We next calculated the ROR-P PRS in the UK Biobank and the Pathways Study (**Figure S3**) and examined the respective associations with breast cancer-specific survival in each study. We expected higher ROR-P PRS to be associated with more aggressive cancers and thus shorter breast cancer-specific survival. In UK Biobank, 66 of 76 SNPs were available for inclusion in the ROR-P PRS. In a Cox proportional hazards regression model adjusted for genetic ancestry, higher ROR-P PRS was associated with worse breast cancer-specific survival (HR per S.D. = 1.12, 95% CI 1.03-1.22, p=0.006) (**Figure 5, Table S7**). Global calibration of the ancestry-adjusted ROR-P PRS in the UK Biobank was acceptable (Gronnesby-Borgan test statistic = 5.8967, p = 0.75) (**Figure S4**).

**Figure 5.**
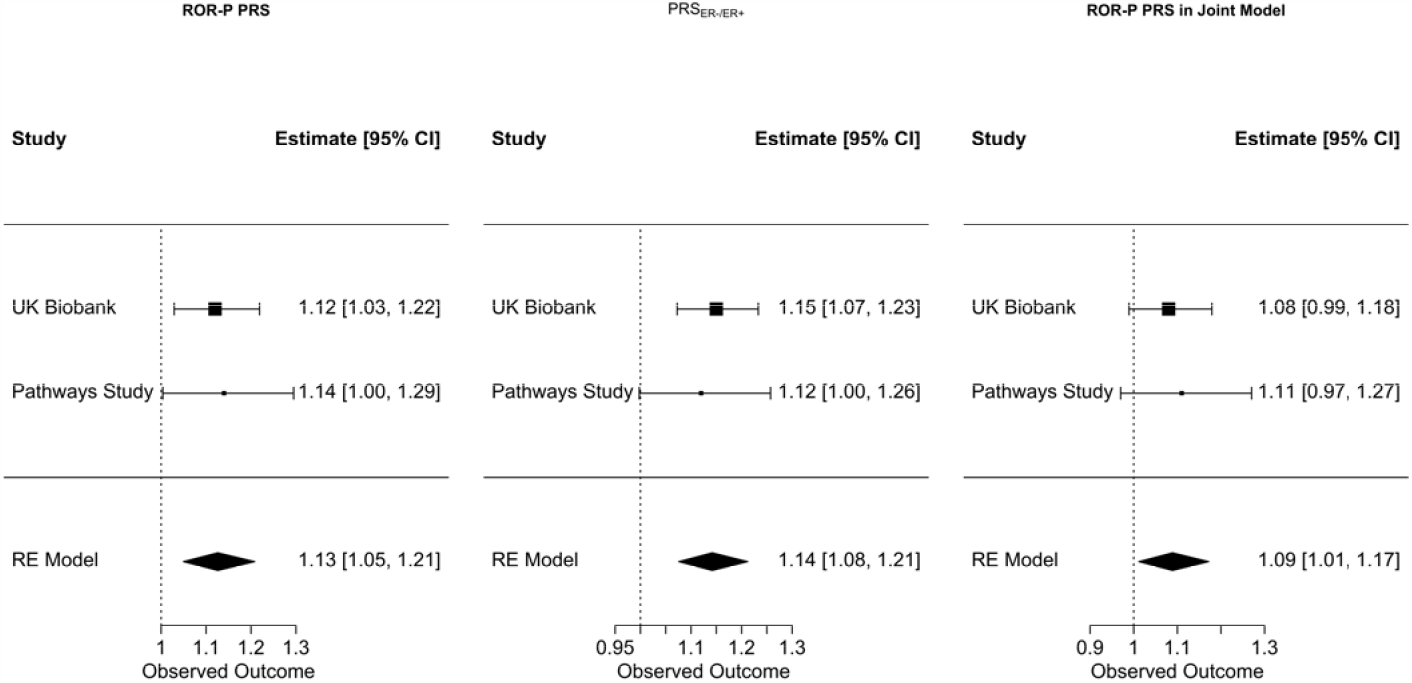
Associations between polygenic risk scores and breast cancer-specific survival. Cox proportional hazards models were constructed for: the polygenic risk score weighted on risk of recurrence (ROR-P PRS), the polygenic risk score for risk of estrogen-negative versus positive breast cancer (ROR-P PRS_ER-/ER+_), and ROR-P PRS adjusted for the effects of PRS_ER-/ER+_ in a joint model. All models were adjusted for genetic ancestry (principal components 1-10). Hazard ratios per standard deviation are shown for UK Biobank, the Pathways Study, and the combined studies using random effects meta-analysis.

In Pathways, 61 of 76 SNPs were available for the PRS. Higher ROR-P PRS was similarly associated with worse survival (HR per S.D. = 1.14, 95% CI 1.00-1.29, p=0.010). In a random-effects meta-analysis of the results from UK Biobank and Pathways, the ROR-P PRS was associated with worse survival (summary HR per S.D. = 1.13, 95% CI 1.05-1.21, p=9.0×10^−4^). No evidence of heterogeneity was found between studies.

Similarly, in Kaplan-Meier analysis of UK Biobank data, the bottom tertile of the ROR-P PRS (corresponding to the lowest predicted ROR-P) was associated with better survival compared with the top and middle tertiles (log-rank chi-squared test statistic = 8.2, p=0.016) (**Figure 6A**). In contrast, the difference in survival between tertiles in the Pathways Study did not reach statistical significance (log-rank chi-squared = 2.8, p=0.25) (**Figure 6B**).

**Figure 6.**
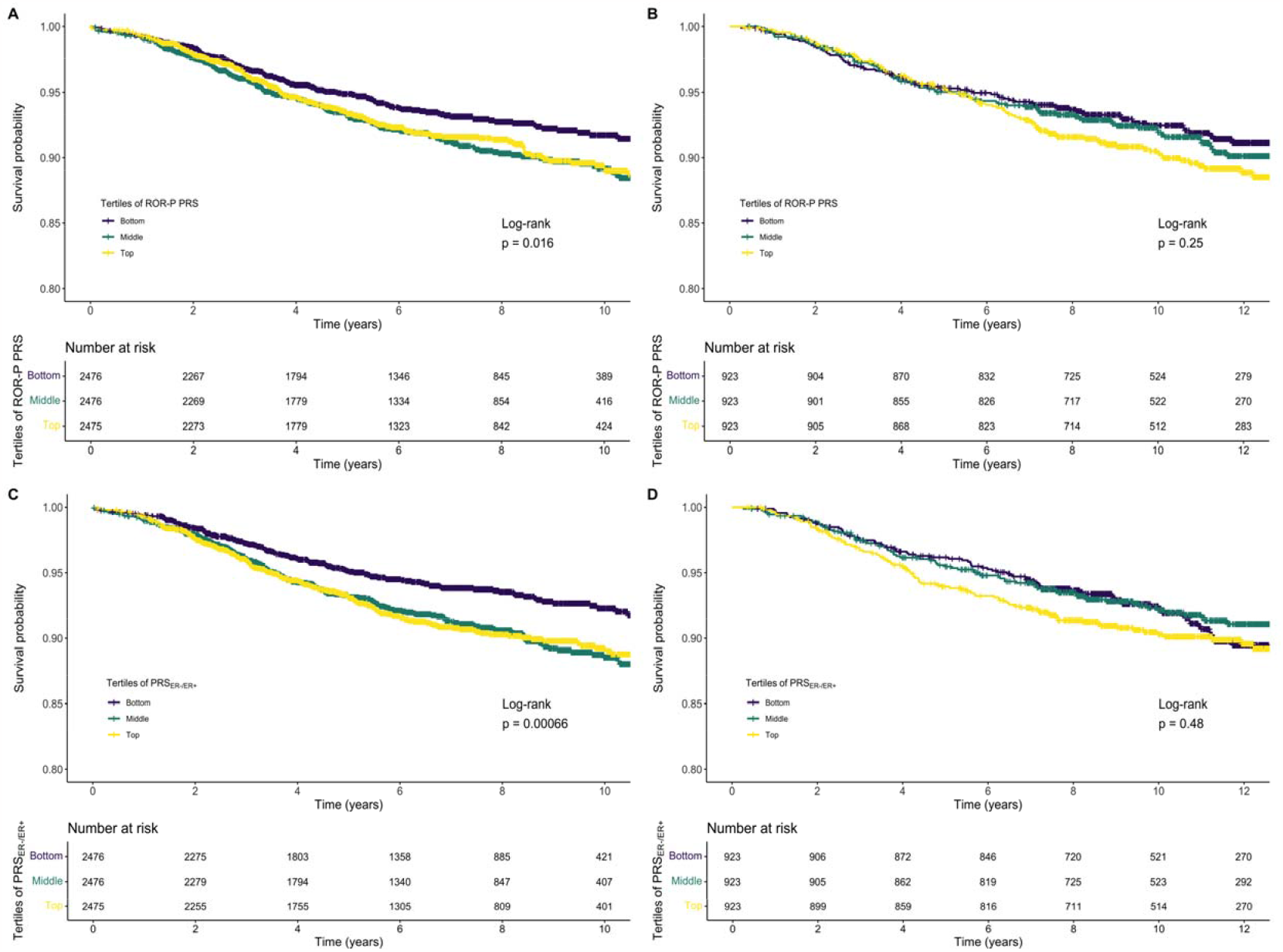
Associations between polygenic risk scores for the risk of recurrence score weighted on proliferation (ROR-P PRS) and risk of estrogen-negative versus positive breast cancer (PRS_ER-/ER+_) versus breast cancer-specific survival. Kaplan-Meier plots of tertiles of the ROR-P PRS in A., the UK Biobank and B., the Pathways Study. Kaplan-Meier plots of tertiles of the PRS_ER-/ER+_ in C., the UK Biobank and D., the Pathways Study. p-values for the log-rank test are shown.

In the Pathways Study, we constructed additional models adjusting for patient-level, tumor, and treatment covariates (**Table S7**). The effect size of the ROR-P PRS did not change with adjustment for age at diagnosis or body mass index. However, its effect was attenuated after including stage at diagnosis (HR per S.D. = 1.11, 95% CI 0.97-1.26, p = 0.12). Including treatment covariates in addition to stage did not substantively change the results. As expected, including measured ROR-P and the ROR-P PRS in the same model attenuated the latter’s effect. Similar results were seen when invasive breast cancer recurrence was used as the outcome in Cox proportional hazards models.

### Evaluation of joint effects with ER-negative PRS and differential effects by ER status

Given the observed association between the ROR-P PRS and ER-negative status as well as the inclusion of ER signaling pathway genes in the ROR-P gene set, we considered the possibility that our ROR-P PRS was recapitulating ER status. Thus, we constructed a comparable PRS for the case-only risk of ER-negative versus ER-positive cancer (PRS_ER-/ER+_) and tested for collinearity and overlapping effects with the ROR-P PRS. In Pathways, we confirmed that the PRS_ER-/ER+_ was associated with increased risk ER-negative versus ER-positive cancer (OR per S.D. = 1.38, 95% CI 1.26-1.52, p=3.1×10^−11^) (**Figure S5**).

In the UK Biobank, higher PRS_ER-/ER+_ was associated with worse breast cancer-specific survival (HR per S.D. = 1.15, 95% CI 1.07-1.23, p=6.8×10^−5^) (**Figure 5**). Kaplan-Meier analysis showed that the bottom tertile of PRS_ER-/ER+_, corresponding to lowest relative risk of ER-negative versus ER-positive cancer, was associated with more favorable survival (**Figure 6C**). In Pathways, there was a similar directional association that did not reach statistical significance (HR per S.D. = 1.12, 95% CI 1.00-1.26, p=0.057) (**Figures 5 & 6D, Table S7**). Meta-analysis of the UK Biobank and Pathways results showed comparable effect sizes between the PRS_ER-/ER+_ and ROR-P PRS (**Figure 5, Table S8**).

We then examined the correlation and joint effects between ROR-P PRS and PRS_ER-/ER+_. There was a modest correlation between the ROR-P PRS and PRS_ER-/ER+_ in UK Biobank and Pathways, Pearson correlation coefficient 0.28, p < 2.2 × 10^−16^, and 0.34, p < 2.2 × 10^−16^, respectively (**Figure S6**). In joint models including both ROR-P PRS and PRS_ER-/ER+_, the effect size of the ROR-P PRS was mildly attenuated (**Table S8, Figure 5**). However, the effect of the ROR-P PRS remained statistically significant in meta-analysis of results from both studies (HR per S.D. = 1.09, 95% CI 1.01-1.17, p=0.012).

To confirm that our ROR-P PRS was not solely recapitulating ER status, we performed additional analyses in the Pathways Study accounting for tumor ER status. Adjusting the ROR-P PRS for ER status led to a mild attenuation of the ROR-P PRS’s effect similar in magnitude to what was observed for the PRS_ER-/ER+_ (**Table S7**). Slight differences in the distributions of the ROR-P PRS were seen in ER-positive versus ER-negative cancers (**Figure S7, Panel A**), with the ROR-P PRS having a stronger effect in ER-positive cancers compared with ER-negative cancers (HR per S.D. = 1.23, 95% CI 1.06-1.43, p = 0.005 versus HR per S.D. = 0.87, 95% CI 0.67-1.12, p = 0.27) (**Figure S7, Panel B**). Taken together, these results strongly suggest the ROR-P PRS contains largely independent information from ER status.

## Discussion

We used associations between breast cancer susceptibility SNPs and tumor gene expression to construct a case-only PRS for ROR-P. The ROR-P PRS was modestly predictive of ROR-P in our development dataset and in an external dataset comprised of tumors from the Pathways Study with measured ROR-P. In survival analysis, higher ROR-P PRS was associated with worse breast cancer-specific survival, with nearly identical effects observed in the UK Biobank and Pathways Study a HR per S.D. of 1.13 (95% CI 1.05-1.21) in meta-analysis. In contrast, PRS_overallBC_ was associated with better survival in the UK Biobank, which is consistent with the results of a large study including nearly 100,000 women with breast cancer^10^. Thus, the associations of the ROR-P PRS and PRS_ER-/ER+_ with worse survival suggest that it is possible to reconfigure PRS to predict aggressive tumors with worse prognosis.

Our findings begin to address an important limitation of current breast cancer PRSs: their preferential associations with less aggressive phenotypes. We decided to fit our PRS to ROR-P, a gene expression-based phenotype, for several reasons. First, standard clinicopathologic markers such as ER status are imperfect proxies for aggressiveness^18,28^. ER-positive cancers display heterogeneous biology and can be divided into luminal A and B subtypes representing low-grade (more indolent) and high-grade (more aggressive) disease, respectively.

Additionally, among molecularly high-risk hormone receptor-positive/HER2-tumors, up to a third are classified as basal^29^. Second, traditional subtyping schemes do not reflect the continuous nature of traits such as receptor expression levels, proliferation, or metastatic potential. Third, aggressiveness is determined by the effects of multiple signaling pathways. For these reasons, continuous multi-gene signatures such as ROR-P may better recapitulate complex, multidimensional traits such as aggressiveness. Prior studies have shown that ROR-P has greater prognostic value than receptor status, grade, and proliferation markers such as Ki67^17^. Fourth, ROR-P has been shown to be heritable^20^, making it an attractive candidate for fitting PRS.

Our targeted approach to PRS building is novel and represents a proof of concept that known breast cancer susceptibility SNPs can be used to fit PRSs to breast cancer aggressiveness. We focused the selection of SNPs for our PRS on variants already known to be associated with breast cancer due to limited power in our datasets for agnostic testing of genome-wide associations with ROR-P and other prognostic features. However, genetic susceptibility to aggressiveness may be influenced by variants outside of those discovered in GWAS for overall breast cancer risk. Prior studies have estimated the heritability of ROR-P to be 13-21% in white women^20^, though it remains to be seen how much of the heritability is explained by known breast cancer susceptibility SNPs versus other common variants. As larger datasets containing germline SNPs and tumor gene expression become increasingly available, it should become feasible to examine a larger pool of candidate SNPs and additional prognostic signatures. With increasing sample size, the use of alternative methods for PRS building such as lasso or elastic net regression^2,30,31^ may also improve prediction. Prognosis may be determined by host factors such as tumor immune microenvironment, and such features could also be used to fit PRS given the role of the germline in shaping immune response^32^.

We also note that a PRS representing the case-case risk of ER-negative versus ER-positive cancer was associated with survival. We included the PRS_ER-/ER+_ as a “positive control” given that ER status has been consistently shown to be prognostic. Whereas the ROR-P PRS and PRS_ER-/ER+_ had comparable magnitudes of association with survival, the more notable finding was the minimal attenuation of the ROR-P PRS’s effects when included in a joint model with PRS_ER-/ER+_. Moreover, the ROR-P PRS had differential associations by ER status with survival, with a stronger effect seen in ER-positive versus ER-negative cancers. This result requires further validation but may reflect the greater heterogeneity in proliferation in ER-positive as opposed to ER-negative cancers, which tend to be more uniformly high-grade. Taken together, our findings suggest that ROR-P PRS captures largely independent prognostic information from ER status, mirroring the composition of the PAM50/ROR-P gene set which includes genes from the estrogen receptor pathway, in addition to genes from multiple others.

A secondary goal of our study was to characterize associations between known breast cancer susceptibility SNPs and ROR-P. The finding that some SNPs are associated with ROR-P is consistent with those of prior studies showing that many of the breast cancer susceptibility SNPs are differentially associated with intrinsic-like subtypes^24,33^. Intrinsic-like subtyping uses immunohistochemical ER, PR and HER2 status plus histologic grade to recapitulate the intrinsic subtypes defined by PAM50, from which ROR-P is derived^18^. Given the moderate correlation between immunohistochemical and expression-based subtyping^34^, we expected to see associations between individual breast cancer SNPs and ROR-P. Our finding of an inverse association between the risk allele and ROR-P for most SNPs adds to the evidence that SNPs discovered in overall breast cancer GWAS are preferentially associated with less aggressive biology^12^.

One strength of this study is our ability to leverage several datasets containing paired germline SNP-tumor gene expression. In particular, the inclusion of samples from the I-SPY 2 TRIAL, which is restricted to molecularly high-risk cancers, allowed us to enrich our PRS development set for aggressive tumors. ROR-P is a widely available signature with strong prognostic value; in one head-to-head comparison, the risk of recurrence (ROR) score was among the highest performing signatures for early and late distant recurrence^35^. Another strength of our study is the use of two independent cohorts to demonstrate the association between ROR-P PRS and survival. We also performed extensive analyses to account for confounding of the ROR-P PRS effect by ER status.

There are several limitations to our study. First, tumor gene expression signatures do not account for spatial or temporal heterogeneity and have imperfect prognostic performance. ROR-P has moderate discrimination for early and late recurrence, with c-statistics of 0.76 and 0.64, respectively^35^. It is possible that fitting the PRS to different prognostic signatures may yield PRSs with stronger associations with prognosis, though these differences would likely be small given the relatively high concordance (approximately 80%) between prognostic signatures^36^. Second, the sample size of our PRS development dataset limited the precision of our effect size estimates for SNPs in our PRS. Thus, the ROR-P PRS may contain some SNPs representing “noise.” Further refinement of the ROR-P PRS in larger datasets is needed, though we are encouraged that the ROR-P PRS displayed consistent effects in both independent validation datasets. Third, our validation was done in breast cancer cases from the UK Biobank and Pathways Study, two studies with fundamentally different designs. Whereas UK Biobank is a population-wide biobanking initiative, Pathways is a breast cancer-specific study that recruited consecutive cases from a single healthcare system in the U.S. The contrasting study designs, settings, and enrollment strategies provide unique strengths and limitations. The robustness of our findings is strengthened by the observation of similar effects across these datasets (considering the smaller sample size of Pathways). However, the UK Biobank did not contain information on breast cancer stage, pathologic features, or treatment, thus limiting our ability to examine the contributions of other determinants of survival and replicate the associations we observed in Pathways between the ROR-P PRS and tumor characteristics. Though we adjusted for confounding by ancestry, it is difficult to rule out residual confounding from other factors, given that we observed a mild attenuation of the ROR-P PRS’s effect in Pathways after accounting for stage and treatment. Fourth, there was limited diversity in the datasets used for ROR-P PRS development and testing. The UK Biobank, despite containing large numbers of breast cancer cases and events, had limited racial and ethnic diversity. Consequently, we restricted Pathways to self-identified White women (representing ∼70% of the study population) for comparability with UK Biobank. Given the disproportionate burden of aggressive cancers in Black/African American women^37^ and Latinas^38^, further work is needed to evaluate and optimize the performance of the ROR-P PRS in diverse populations.

We believe that the potential clinical utility of the ROR-P PRS could lie in enhancing risk stratification for screening and prevention. Ongoing trials of risk-based screening are using models for overall breast cancer risk to assign screening recommendations^5^, but these models do not account for the risk of developing aggressive cancer. Aggressive cancers are over-represented in younger women, including those younger than the starting age for initiating screening recommended by current clinical guidelines. Thus, the ROR-P PRS, PRS_ER-/ER+_, and similar PRS could be tested as modifiers to established risk models, particularly to identify women who should be offered screening at an earlier age, at shorter intervals, or using high-sensitivity modalities such as magnetic resonance imaging. Women at elevated risk of aggressive cancers may be ideal candidates for prevention and interception trials. In addition, it has been suggested that PRS predictive of cancer-related death may be more valuable than PRS for overall risk in selecting individuals with the greatest net benefit from screening – particularly for cancers (such as breast and prostate) where overdiagnosis is common^39^.

Our work represents a first step toward the prediction of phenotype-specific breast cancer risk. Future studies should seek to refine the methods used to construct PRS for aggressiveness and evaluate the performance of these PRSs in diverse populations with larger numbers of non-White individuals. Since PRS development and testing were done using a case-only design, further work is needed to examine case-control associations and to test the performance (e.g. discrimination, calibration, net reclassification improvement) of PRSs in predicting aggressive, poor-prognosis cancers on a population basis. Concurrent efforts should seek to refine the understanding of how the germline contributes to gene expression and other somatic features, and how these relationships shape tumor aggressiveness. Such analyses should become increasingly feasible with the growing availability of integrated datasets containing germline and somatic genomic data.

## Methods

### Study population

The PRS development phase of our study included invasive breast cancers from three datasets with paired germline SNP-tumor gene expression data: the Cancer Genome Atlas (TCGA)^21^, a publicly available pan-cancer atlas; Molecular Taxonomy of Breast Cancer International Consortium (METABRIC)^22^, a breast cancer genomic profiling study, and the Investigation of Serial Studies to Predict Your Therapeutic Response with Imaging And moLecular analysis 2 (I-SPY 2) TRIAL (NCT01042379)^23^ (**Table S1**). The I-SPY 2 TRIAL is an ongoing clinical trial comparing multiple novel therapeutic agents for neoadjuvant treatment of locally advanced breast cancer. All cancers in I-SPY 2 must be considered molecularly high-risk according to clinicopathologic data or results of the MammaPrint prognostic signature^40^; as a result, all tumors undergo gene expression profiling. Cases were included in our analysis based on the following criteria: In TCGA, we included invasive breast cancers (n=953); samples corresponding to the primary tumor were included while those corresponding to recurrences or normal tissue were excluded. In METABRIC, we included cases with available SNP genotyping data (n=496). In I-SPY 2, genotyping has been completed for the first 1,400 cases; of these, a subset had available ROR-P calls (n=914).

To test the association between ROR-P PRS and clinical outcomes, we analyzed participants from two studies containing longitudinal follow-up of women diagnosed with breast cancer, the UK Biobank and the Pathways Study (**Table S2**). UK Biobank is a population-based cohort that enrolled individuals aged 40-69 years across the UK between 2006-2010, with cancer diagnoses and deaths ascertained from national registries^41^. To identify women with breast cancer, we used International Classification of Diseases (ICD) codes: ICD-9 (175, 1740, 1741, 1742, 1743, 1744, 1745, 1746, 1748, 1749, and 2330) and ICD-10 (C500, C501, C502, C503, C504, C505, C506, C508, C509, D050, D051, D057, and D059). We converted all codes to ICD-10-CM, then ICD-0-3, using Surveillance, Epidemiology, and Endpoints Registry (SEER) conversion tables. We then linked these ICD codes to SEER site recodes (2008). The SEER site recode 26000 was used for breast cancer. We defined breast cancer-related deaths as those for which breast cancer was indicated as contributing to the death (ICD-10 code C509). For non-censored individuals, the date of last follow-up was December 31, 2019. Given the ICD codes used to identify breast cancer patients included those pertaining to invasive and in situ disease, we created indicator variables for these categories. We restricted our analysis to self-identified British White women with incident invasive breast cancer (n = 7,427) to mitigate potential biases related to survivorship and temporal treatment trends. Incident cancers were those with a diagnosis date occurring after the date of UK Biobank enrollment.

The Pathways Study is a longitudinal cohort of women diagnosed with breast cancer at Kaiser Permanente Northern California. Participants included women aged 21 years and older with a first diagnosis of invasive breast cancer between 2006-2013^42^. Data on treatment, recurrence, and death were ascertained from the Kaiser Permanente Northern California Cancer Registry, as well as electronic medical records. Imputed genotype data was available for 3,973 of 4,377 total participants. For comparability with UK Biobank, we included participants with compete clinical and genetic data who were of self-reported White race (n = 2,769)^43^.

The pooled analysis described in this manuscript was approved by the Biomedical Research Alliance of New York.

### Genotyping

We performed SNP genotyping using array-based methods and imputed genotypes to population-based references (**Tables S1 and S2**). We estimated genetic ancestry by generating the first 10 principal components (PCs) based on genotyped markers using Plink (version 1.9). SNPs with >5% missingness were excluded. SNPs with <5% missingness were randomly assigned a genotype weighted on the distribution of genotypes calculated by the Hardy-Weinberg equation. This calculation used allele frequencies for the respective SNP among individuals without missing genotypes.

### Tumor gene expression

Tumor gene expression was measured on the platforms detailed in **Table S1**. ROR-P is the composite of ROR-S, the linear combination of PAM50 subtype-centroid correlations, and a proliferation score calculated from an 11-gene subset of the PAM50 gene set. To generate ROR-P calls, we first performed batch correction of raw gene expression levels (R package *comBAT*)^44^. For genes with multiple probes, we collapsed expression levels to the mean across all probes for the gene. Performing PAM50/ROR-P calls on a batch of tumors requires the target dataset to have a similar distribution of ER-positive and ER-negative cases to the original PAM50 training set. To address this “population assumption,” we created a subsample including all ER-cancers plus an equal number of randomly selected ER+ cases. We repeated the subsampling procedure 1000 times and calculated for each repetition the median expression of each PAM50 gene. For each gene, we calculated the median of the 1000 medians and subtracted it from the collapsed expression levels. We then used these normalized expression levels to calculate ROR-P as previously described^18^.

### Construction of PRS for ROR-P

We tested 226 candidate SNPs with genome-wide significant associations (p<5×10^−8^) in prior genome-wide association studies (GWAS) of overall breast cancer susceptibility^1,45,46^, or a related phenotype such as ER-negative^13,47^ or intrinsic-like subtype^33^, age of onset^25^, or prognosis/survival^48-52^ (**Table S4**). We identified candidate SNPs and obtained summary statistics from the Breast Cancer Association Consortium (BCAC)^1^, accessed at https://bcac.ccge.medschl.cam.ac.uk/bcacdata/oncoarray/oncoarray-and-combined-summary-result/gwas-summary-results-breast-cancer-risk-2017, and the GWAS Catalog^53^. We started with 271 SNPs and performed linkage disequilibrium (LD) clumping using the LDLink (R package *LDlinkR*)^54^. Within pairs of SNPs in LD (r^2^ ≥ 0.2 in European populations), we kept the SNP with the lower published p-value for association with breast cancer susceptibility. After LD pruning, 226 SNPs remained.

To address potential confounding, we constructed the following linear regression model within pooled TCGA, METABRIC, and I-SPY 2 data:

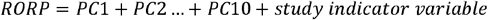

We then regressed the model residual against each individual SNP and obtained the β coefficients and p-values for each association.

To identify the best performing model for ROR-P, we constructed PRSs according to varying p-value thresholds (0.025-0.5) for SNP inclusion. We used 5-fold cross-validation (R package *caret*)^55^ to estimate the r^2^ of each PRS against the residual in the leave-out subset. We repeated this process 10 times. After identifying the p-value threshold with the highest r^2^, we included in our PRS all SNPs with a p-value below this threshold. Finally, we obtained within the overall dataset the coefficients of individual SNP associations with ROR-P, adjusted for genetic ancestry and an indicator variable for study.

To calculate the ROR-P PRS in UK Biobank and Pathways, we applied the SNP coefficients derived from our PRS development set to the genotype data, coded as risk allele dosage. Specifically, the PRS was calculated as:

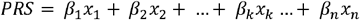

Where *β*_*k*_ is the per-allele linear regression coefficient for ROR-P associated with SNP *k, x*_*k*_ is the genotype coded as number of risk alleles present, and *n* is the total number of SNPs in the PRS.

### Construction of PRS for overall and ER-specific cancer risk

We generated two comparator PRSs and tested their associations with breast cancer-specific survival: 1) a 274-SNP PRS for overall risk of breast cancer (UK Biobank only), PRS_overallBC_; and 2) a 205-SNP PRS representing the case-case relative risk of developing ER-negative versus ER-positive disease, PRS_ER-/ER+_. PRS_overallBC_ was derived from a published 313-SNP PRS for overall breast cancer^2^. In UK Biobank, we were able to retrieve genotypes for 274 of the 313 SNPs in the PRS. The candidate SNPs for the PRS_ER-/ER+_ were the same 271 SNPs considered for inclusion in the ROR-P PRS. The effect sizes of the SNPs were taken from summary statistics for associations with ER-negative and ER-positive breast cancer, as reported in the BCAC iCOGs and OncoArray studies (https://gwas.mrcieu.ac.uk/, ieu-a-1127 and ieu-a-1128, respectively)^56^. We used the β coefficients calculated from meta-analysis of these studies. Given the β coefficients for ER-negative and ER-positive cancers were derived from case-control comparisons with the same control group, we subtracted the β coefficient for ER-positive cancer from the β coefficient for ER-negative cancer^2^ to obtain the case-case β coefficient for ER-negative versus ER-positive risk. Summary statistics were available for 237 SNPs. After LD pruning, 205 SNPs remained, of which 193 had available genotypes in UK Biobank and 189 had available genotypes in the Pathways Study. We calculated the PRS using a previously described method^3,57^.

### Statistical analysis

As a form of external validation, we examined associations between ROR-P PRS and tumor features such as ER status, HER2 status, histologic grade, and intrinsic-like subtype in the Pathways Study. For binary features such as ER and HER status, we used t-tests to compare mean ROR-P PRS between categories. We also constructed logistic regression models adjusted for genetic ancestry principal components 1-10. (PC1-PC10). For categorical outcomes with three or more categories, such as grade and intrinsic-like subtype, we used analysis of variance (ANOVA) tests and constructed multinomial logistic regression models adjusted for genetic ancestry. We also estimated the correlation between ROR-P PRS and actual ROR-P using the Pearson correlation coefficient in the subset of tumors that had undergone gene expression profiling.

To confirm past findings that a PRS representing overall breast cancer risk was associated with improved survival, we first performed survival analysis of PRS_overallBC_ in the UK Biobank. We constructed a Cox proportional hazards regression model with PRS_overallBC_ as the predictor and genetic ancestry PC1-PC10 as covariates. We normalized PRS_overallBC_ to the mean and standard deviation among cases. We also used Kaplan-Meier survival analysis (R packages *survival* and *survminer*)^58^ to examine the association between tertiles of PRS_overallBC_ and breast cancer-specific survival and tested for differences between tertiles using log-rank tests.

To examine the respective associations between ROR-P PRS and PRS_ER-/ER+_ and breast cancer-specific survival in UK Biobank and Pathways, we constructed Cox proportional hazards models for each PRS, with adjustment for genetic ancestry as above. We normalized the ROR-P PRS to the mean and log-normalized the PRS_ER-/ER+_. We also performed Kaplan-Meier survival analysis using tertiles of the ROR-P PRS and PRS_ER-/ER+_. To examine joint effects between ROR-P PRS and PRS_ER-/ER+_, we calculated the Pearson correlation coefficient between the two PRSs. We also constructed Cox proportional hazards models including terms for ROR-P PRS and PRS_ER-/ER+_. To synthesize the results of Cox proportional hazards models from UK Biobank and Pathways, we performed random-effects meta-analysis using a restricted maximum likelihood estimator (R package *metafor*)^59^. We evaluated for heterogeneity between studies using Cochran’s Q test and calculated the I^2^ index.

We evaluated calibration of the Cox model containing ancestry-adjusted ROR-P PRS using the UK Biobank data. We obtained bias-corrected estimates of predicted versus observed breast cancer-specific survival at 5 years using boostrapping with 200 repeats (R package *rms*). We compared the predicted survival probabilities for 10 evenly divided strata of risk versus the Kaplan-Meier “observed” estimates for each stratum. We also performed the Gronnesby-Borgan goodness-of-fit test for the Cox model (R package *survMisc*).

In Pathways, we built additional nested ROR-P PRS models adjusted for the following combinations of covariates: age at diagnosis and body mass index (Model 2); age at diagnosis, body mass index, and stage at diagnosis (Model 3); age at diagnosis, body mass index, stage at diagnosis, and binary variables corresponding to receipt of the following treatments: radiation therapy, chemotherapy, trastuzumab, and hormone therapy (Model 4); measured ROR-P (Model 5); and ER status (Model 6). Lastly, we constructed nested models containing ROR-P PRS and the same combinations of covariates as Models 2-6 but using invasive breast cancer recurrence as the outcome. We also examined differential effects of the ROR-P PRS by ER status by constructing separate Cox proportional hazards models for ER-positive and ER-negative cancers.

All statistical tests were two-sided with α = 0.05. Analyses were done using R version 4.1.2 (R Foundation, Vienna, Austria).

### Code availability

Statistical code used to conduct the analyses can be found at https://github.com/shiehy/ROR-P_PRS.

## Data Availability

The Cancer Genome Atlas data are publicly available and were accessed through the Broad GDAC Firehose (https://gdac.broadinstitute.org/). METABRIC data are available upon application to the METABRIC data access committee and were accessed through to the European Genome-Phenome Archive (https://ega-archive.org). I-SPY 2 data are available upon application to the I-SPY 2 TRIAL Data Access Committee. The UK Biobank data are available to approved researchers registered with the UK Biobank. The research was conducted with approved access to UK Biobank data under application number 14105. The Pathways Study genotype data are available on The database of Genotypes and Phenotypes (dbGaP) under study accession phs001534.v1.p1.

Clinical and outcomes data are available upon application to the Pathways Study Steering Committee.

## Data Availability

All data produced in the present study are available upon reasonable request to the authors

## Acknowledgements

This work was supported by funding from the National Cancer Institute (K08 CA237829, K24 CA169004, U01 CA196406, P01 CA210961, R01 CA105274, U01 CA195565, R01 CA129059) and the National Human Genome Research Institute (X01 HG008335). The I-SPY 2 Study was also supported by the Quantum Leap Healthcare Collaborative and the Foundation for the National Institutes of Health. The results shown here are in whole or part based upon data generated by the TCGA Research Network: https://www.cancer.gov/tcga.

## Author Contributions

Study conception and design: Y.S. and E.Z. Data abstraction and acquisition: Y.S., C.Y., D.M.W., G.L.H., L.B.S., S.H., J.L.N., P.M., L.K., Q.Z., S.Y. Statistical analysis: Y.S., J.M.R., C.Y., D.M.W., D.H., and R.S.H. Interpretation of data: Y.S., C.Y., D.M.W., L.H.K., and E.Z. Manuscript preparation: Y.S. and E.Z. Manuscript editing and critical review: all authors. Financial support, resources, and study supervision: Y.S., M.L.K., B.J.C., J.S.W., L.H.K., L.V.V., L.J.E., and E.Z.

## Materials & Correspondence

Correspondence to Yiwey Shieh.

