## Supplementary tables and figures for "Development and testing of a polygenic risk score for breast cancer aggressiveness"

**Supplementary Materials**

**Table S1. Datasets used in the development of the polygenic risk score for risk of recurrence score weighted on proliferation (ROR-P PRS)**

| **Study** | **Number of cases** | **Selection criteria for cases** | **Gene expression profiling method** | **SNP genotyping platform** | **Imputation reference** | **Quality control** |
| --- | --- | --- | --- | --- | --- | --- |
| The Cancer Genome Atlas (TCGA) | 953 | Invasive breast cancers, recurrences excluded | RNA-Seq | Affymetrix SNP 6.0 | 1000 Genomes | SNPs excluded if minor allele frequency <0.5% or variant missing rate >5%. Individuals excluded if missing call rate > 5%. |
| Molecular Taxonomy of Breast Cancer International Consortium (METABRIC) | 496 | Genotyped subset | Illumina HT-12 beadchip | Affymetrix SNP 6.0 | 1000 Genomes |  |
| Investigation of Serial Studies to Predict Your Therapeutic Response with Imaging And molecular analysis 2 (I-SPY 2 TRIAL) | 914 | First 1400 consecutive cases underwent genotyping; ROR-P available for a subset | Agilent 44k array | Affymetrix Axiom Precision Medicine Diversity Array | 1000 Genomes |  |

**Table S2. Datasets used to test the association between the polygenic risk score for the risk of recurrence score weighted on proliferation (ROR-P PRS) with clinical outcomes**

| **Study** | **Cases** | **Breast cancer deaths** | **Years of diagnosis, cases** | **Length of follow-up, median (IQR)** | **SNP genotyping platform** | **Imputation reference** | **Quality control** |
| --- | --- | --- | --- | --- | --- | --- | --- |
| UK Biobank | 7,427 | 544 | 2006-2019 | 6.4 years | Affymetrix UK Biobank Axiom Array | Haplotype Reference Consortium (primary); UK10K, 1000 Genomes (secondary) | SNPs excluded if: variant missing rate >3%; minor allele frequency <0.01; heterozygosity more than 5 standard deviations from mean |
| Pathways Study | 2,769 | 241 | 2006-2013 | 10.7 years | Illumina MultiEthnic Global-8 | Haplotype Reference Consortium | SNPs excluded if: variant missing rate >2%; >1 Mendelian error in the HapMap trios included in genotyping; Hardy–Weinberg equilibrium (p < 1×10^−4^); discordance between duplicate samples; or positional duplicated variants. |

**Table S3. Characteristics of studies used for survival analysis of the polygenic risk score for the risk of recurrence score weighted on proliferation (ROR-P PRS)**

|  | **UK Biobank** | **Pathways Study** |
| --- | --- | --- |
| Characteristic | N = 7,427^1^ | N = 2,769^1^ |
| Age at diagnosis (years) | 64 (57, 69) | 62 (54, 70) |
| Breast cancer deaths | 544 (7.3%) | 241 (8.7%) |
| Follow-up time (years) | 6.4 (3.7, 9.1) | 10.7 (8.2, 12.3) |
| Body mass index |  | 27 (24, 32) |
| Unknown |  | 1 |
| Estrogen receptor status |  |  |
| positive |  | 2,386 (86%) |
| negative |  | 382 (14%) |
| Unknown |  | 1 |
| HER2 status |  |  |
| positive |  | 319 (12%) |
| negative |  | 2,337 (88%) |
| Unknown |  | 113 |
| Histologic grade |  |  |
| 1 |  | 795 (31%) |
| 2 |  | 1,192 (46%) |
| 3 |  | 612 (24%) |
| Unknown |  | 170 |
| IHC subtype |  |  |
| Luminal A |  | 2,066 (78%) |
| Luminal B |  | 224 (8.4%) |
| Basal |  | 270 (10%) |
| HER2 |  | 95 (3.6%) |
| Unknown |  | 114 |
| ROR-P |  | 45 (17, 63) |
| Unknown |  | 2,285 |
| ROR-P Group |  |  |
| Low |  | 96 (20%) |
| Medium |  | 194 (40%) |
| High |  | 194 (40%) |
| Unknown |  | 2,285 |
| ^1^Median (IQR); n (%)  HER2, human epidermal growth factor receptor 2; IQR, interquartile range; ROR-P, risk of recurrence score weighted on proliferation | | |

**Table S4. Candidate single nucleotide polymorphisms for polygenic risk score for the risk of recurrence score weighted on proliferation (ROR-P PRS)**

See separate Excel file.

| **p-value threshold** | **Number of SNPs** | **RMSE** | **R-squared** | **MAE** |
| --- | --- | --- | --- | --- |
| 0.6 | 124 | 21.75565 | 0.0398811 | 17.72236 |
| 0.575 | 119 | 21.70951 | 0.0416786 | 17.68096 |
| 0.55 | 111 | 21.66005 | 0.0433478 | 17.65135 |
| 0.525 | 107 | 21.6579 | 0.0425431 | 17.64337 |
| 0.5 | 104 | 21.63518 | 0.043404 | 17.62402 |
| 0.475 | 98 | 21.59115 | 0.0449304 | 17.58716 |
| 0.45 | 92 | 21.5553 | 0.0465036 | 17.56158 |
| 0.425 | 91 | 21.55257 | 0.0465777 | 17.56454 |
| 0.4 | 84 | 21.49866 | 0.0490263 | 17.51114 |
| 0.375 | 76 | 21.44928 | 0.0509436 | 17.48951 |
| 0.35 | 67 | 21.52258 | 0.0431191 | 17.53252 |
| 0.325 | 62 | 21.50549 | 0.0431755 | 17.52775 |
| 0.3 | 59 | 21.63901 | 0.0322063 | 17.6672 |
| 0.275 | 53 | 21.6204 | 0.0322852 | 17.64313 |
| 0.25 | 50 | 21.59195 | 0.0337823 | 17.61451 |
| 0.225 | 48 | 21.58708 | 0.0336946 | 17.61529 |
| 0.2 | 42 | 21.59427 | 0.0320937 | 17.61333 |
| 0.175 | 34 | 21.59588 | 0.0304884 | 17.62185 |
| 0.15 | 28 | 21.63223 | 0.0259714 | 17.643 |
| 0.125 | 23 | 21.65455 | 0.0233348 | 17.66418 |
| 0.1 | 19 | 21.68273 | 0.0198944 | 17.68723 |

**Table S5. Performance of polygenic risk scores for the risk of recurrence score weighted on proliferation (ROR-P PRS) by p-value threshold**

**Table S6. Associations between single nucleotide polymorphisms (SNPs) and the risk of recurrence score weighted on proliferation (ROR-P) for the 76-SNP ROR-P polygenic risk score**

See separate Excel file.

**Table S7. Results of Cox proportional hazards models of the polygenic risk score for the risk of recurrence score weighted on proliferation (ROR-P PRS) versus breast cancer survival and invasive recurrence in the Pathways Study**

|  | Breast cancer death | | Invasive recurrence | |
| --- | --- | --- | --- | --- |
|  | HR (95% CI) | p-value | HR (95% CI) | p-value |
| Model 1^a^  ROR-P PRS | 1.14 (1.01-1.30) | 3.8x10^-2^ | 1.11 (1.00-1.24) | 4.1x10^-2^ |
| Model 2^a^  ROR-P PRS  Age at diagnosis  BMI | 1.14 (1.00-1.29)  1.01 (1.00-1.02)  1.02 (1.00-1.04) | 4.3x10^-2^  4.8x10^-2^  1.6x10^-2^ | 1.12 (1.01-1.24)  1.00 (0.99-1.01)  1.01 (0.99-1.02) | 3.9x10^-2^  4.0x10^-1^  3.8x10^-1^ |
| Model 3^a,b^  ROR-P PRS  Age at diagnosis  BMI  Stage^c^  I  II  III  IV | 1.11 (0.97-1.26)  1.02 (1.01-1.03)  1.02 (1.00-1.04)  Ref.  2.65 (1.88-3.73)  8.56 (5.94-12.3)  44.5 (28.1-70.4) | 1.2x10^-1^  2.5x10^-3^  6.6x10^-2^  3.0x10^-8^  <2x10^-16^  <2x10^-16^ | 1.11 (1.00-1.23)  1.00 (0.99-1.01)  1.00 (0.99-1.02)  Ref.  2.30 (1.79-2.97)  5.44 (4.06-7.28)  5.43 (3.04-9.69) | 5.3x10^-2^  6.2x10^-1^  9.0x10^-1^  1.2x10^-10^  <2x10^-16^  1.0x10^-8^ |
| Model 4^a,b^  ROR-P PRS  Age at diagnosis  BMI  Stage^c^  I  II  III  IV  Chemotherapy  Taxane therapy  Hormonal therapy  Radiation therapy  Trastuzumab | 1.10 (0.96-1.24)  1.03 (1.02-1.04)  1.02 (1.00-1.04)  Ref.  1.95 (1.33-2.84)  5.90 (3.87-8.99)  36.2 (22.4-58.4)  2.15 (1.32-3.50)  0.91 (0.58-1.45)  0.50 (0.37-0.66)  0.74 (0.55-1.00)  0.53 (0.37-0.75) | 1.6x10^-1^  8.6x10^-6^  7.1x10^-2^  5.5x10^-4^  <2x10^-16^  <2x10^-16^  2.0x10^-3^  7.0x10^-1^  1.3x10^-6^  5.0x10^-2^  4.5x10^-4^ | 1.10 (0.99-1.22)  1.01 (1.00-1.02)  1.00 (0.99-1.02)  Ref.  2.08 (1.57-2.75)  4.75 (3.39-6.64)  4.38 (2.40-7.97)  1.26 (0.83-1.91)  0.98 (0.65-1.47)  0.73 (0.58-0.91)  0.88 (0.69-1.12)  0.89 (0.68-1.18) | 6.7x10^-2^  2.7x10^-1^  8.6x10^-1^  2.8x10^-7^  <2x10^-16^  1.4x10^-6^  2.8x10^-1^  9.1x10^-1^  5.7x10^-3^  2.9x10^-1^  4.2x10^-1^ |
| Model 5^a, c^  ROR-P PRS  ROR-P | 0.95 (0.77-1.18)  1.02 (1.01-1.03) | 6.5x10^-1^  9.8x10^-4^ | 1.01 (0.83-1.23) 1.01 (1.00-1.02) | 9.1x10^-1^  4.8x10^-2^ |
| Model 6^a^  ROR-P PRS  ER status  Positive  Negative | 1.12 (0.99-1.27)  Ref.  2.47 (1.85-3.30) | 8.2x10^-2^  7.3x10^-10^ | 1.10 (0.99-1.23)  Ref.  1.79 (1.38-2.32) | 6.3x10^-2^  1.3x10^-5^ |

^a^All models are adjusted for genetic ancestry, PC1-10

^b^Model includes subset of 2,689 cancers with available stage data with 230 breast cancer deaths

^c^American Joint Committee on Cancer (AJCC) Cancer Staging Manual, 6^th^ Edition

^c^Model includes subset of 484 cancers with available ROR-P with 87 events

Abbreviations: BMI, body mass index; CI, confidence interval; ER, estrogen receptor; HR, hazard ratio; PRS, polygenic risk score; ROR-P, risk of recurrence score weighted on proliferation; Tx., therapy

**Table S8. Results of Cox proportional hazards models for the risk of recurrence score weighted on proliferation (ROR-P PRS), risk of estrogen-negative versus positive breast cancer (PRS_ER-/ER+_), and joint model including both ROR-P PRS and PRS_ER-/ER+_**

|  | UK Biobank | | Pathways | | Meta-analysis | | | | |
| --- | --- | --- | --- | --- | --- | --- | --- | --- | --- |
|  | HR (95% CI) | p-value | HR (95% CI) | p-value | HR (95% CI) | p-value | Q | P_het_ | I^2^ |
| ROR-P PRS | 1.12 (1.03-1.22) | 6.2x10^-3^ | 1.14 (1.00-1.29) | 1.0x10^-2^ | 1.13 (1.05-1.21) | 1.0x10^-3^ | 0.05 | 0.82 | 0% |
| PRS_ER-/ER+_ | 1.15 (1.07-1.23) | 6.8x10^-5^ | 1.12 (1.00-1.26) | 5.7x10^-2^ | 1.14 (1.08-1.22) | <1.0x10^-4^ | 0.12 | 0.73 | 0% |
| Joint model  ROR-P PRS  PRS_ER-/ER+_ | 1.08 (0.99-1.18)  1.13 (1.05-1.21) | 6.7x10^-2^  1.0x10^-3^ | 1.11 (0.97-1.27)  1.08 (0.96-1.23) | 1.2x10^-1^  2.0x10^-1^ | 1.10 (1.02-1.18)  1.12 (1.05-1.19) | 1.4x10^-2^  4.0x10^-4^ | 0.33  0.39 | 0.57  0.53 | 0%  0% |

All models are adjusted for genetic ancestry, PC1-10

Abbreviations: CI, confidence interval; HR, hazard ratio; PRS, polygenic risk score; ROR-P, risk of recurrence score weighted on proliferation

**Figure S1. Distribution of the risk of recurrence score weighted on proliferation (ROR-P) in the polygenic risk score (PRS) development datasets**

Histograms of ROR-P in breast cancers from (A) The Cancer Genome Atlas, n=953; (B) Molecular Taxonomy of Breast Cancer International Consortium (METABRIC), n=496; and (C) Investigation of Serial Studies to Predict Your Therapeutic Response with Imaging And molecular analysis 2 (I-SPY 2 TRIAL), n=914.

A.

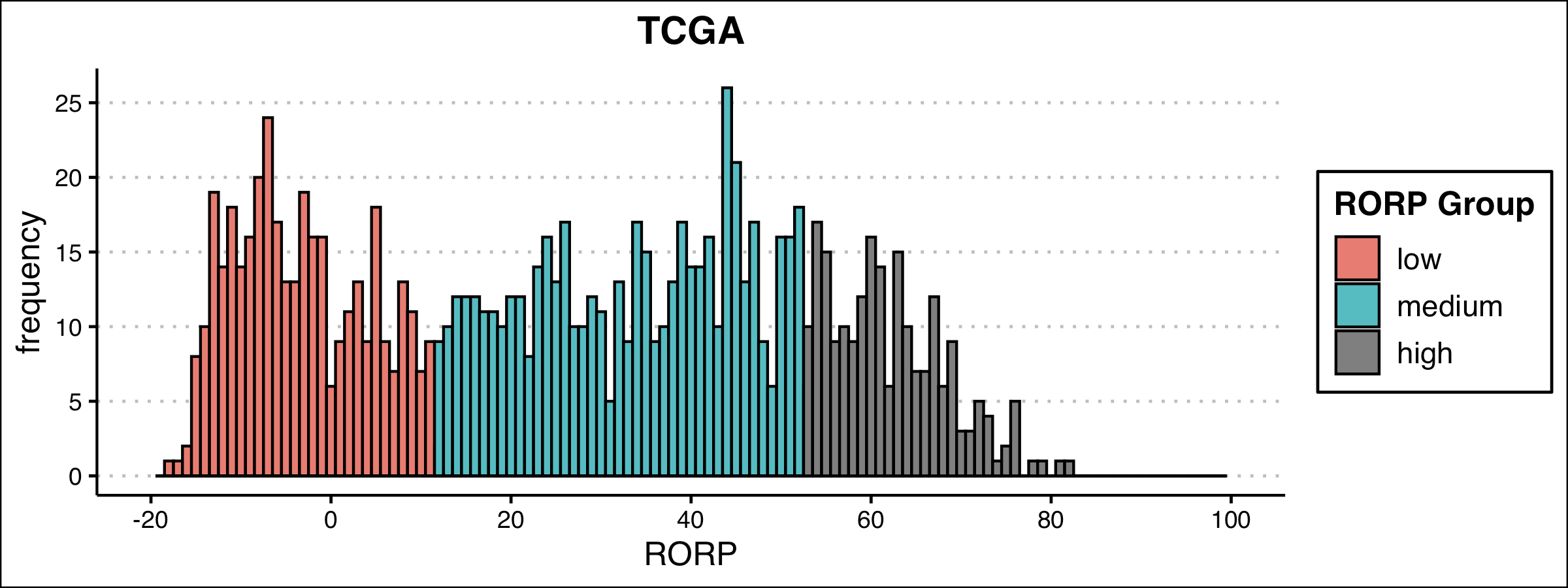

B.

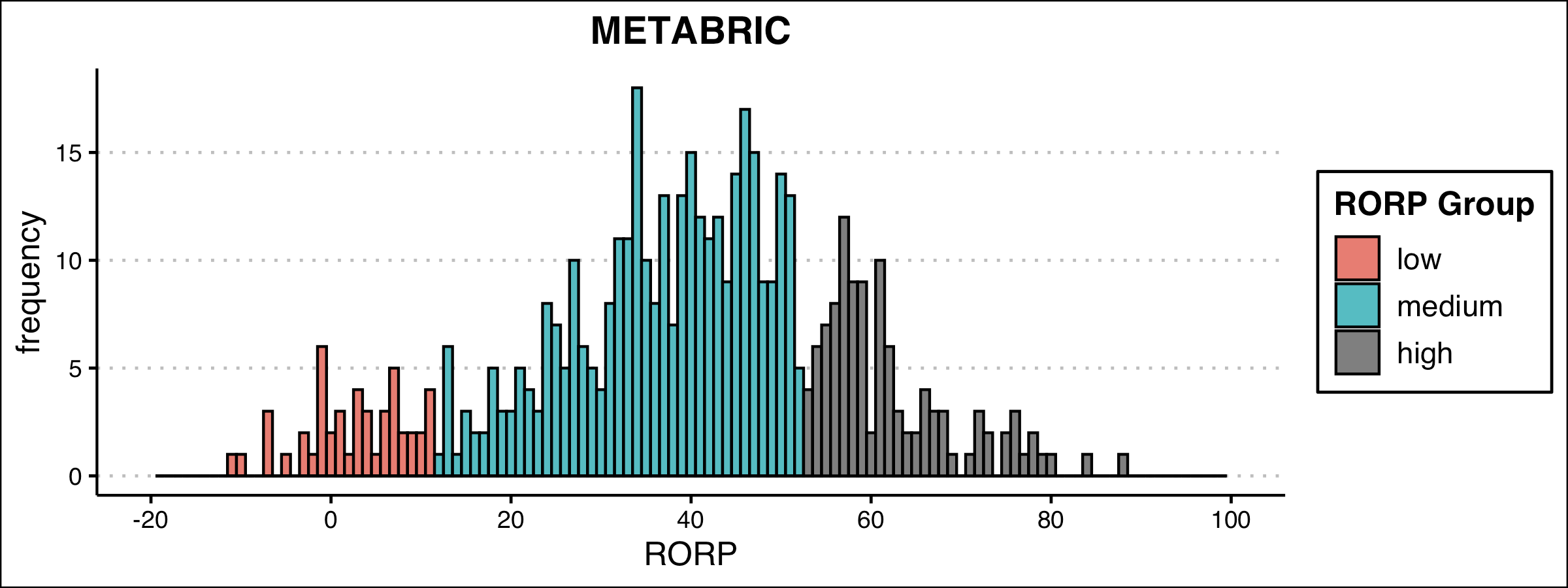

C.

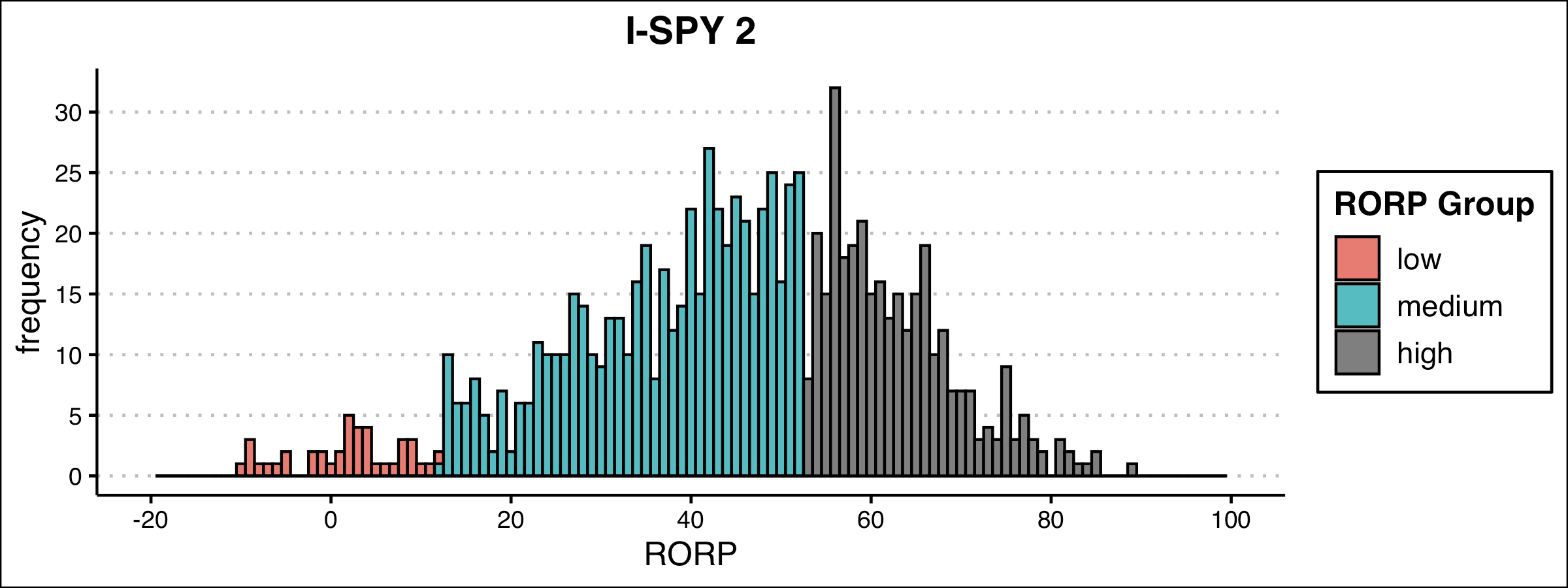

**Figure S2. Distribution of the risk of recurrence score weighted on proliferation (ROR-P) in the Pathways Study**

Histogram of ROR-P breast cancers from the Pathways Study that underwent tumor gene expression profiling, n=484.

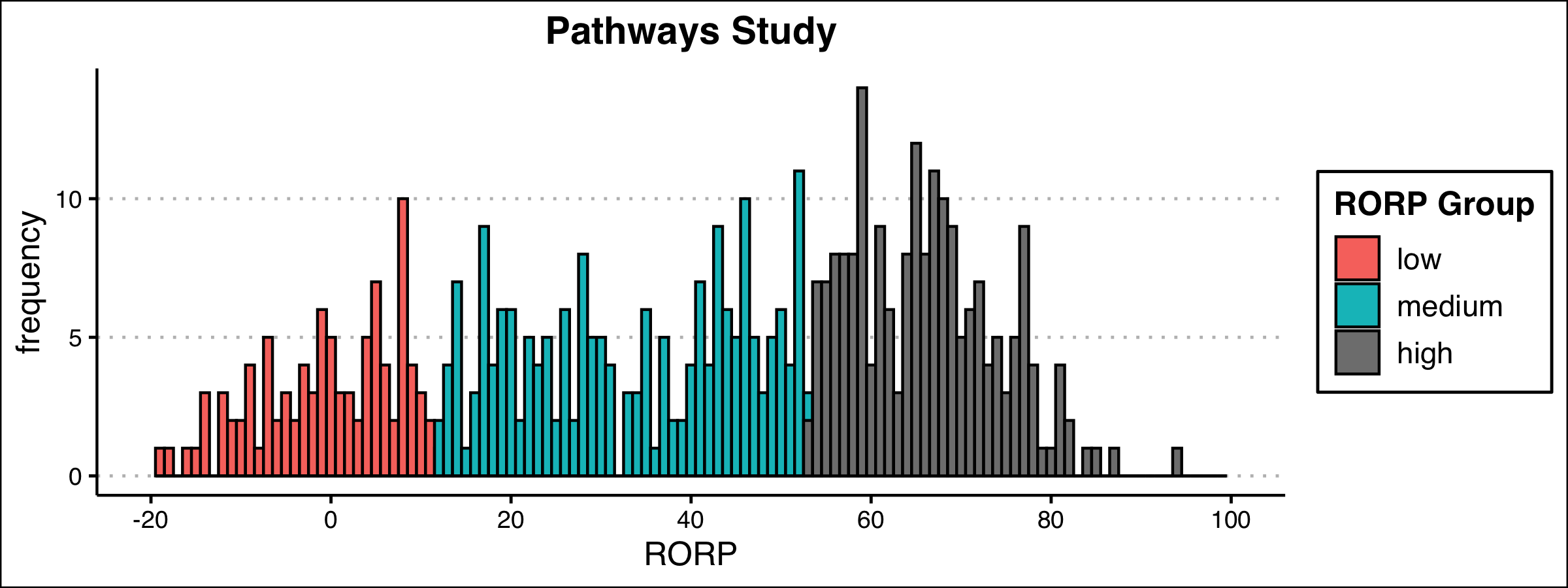

**Figure S3. Distributions of polygenic risk score for the risk of recurrence score weighted on proliferation (ROR-P PRS) in UK Biobank and the Pathways Study**

Histograms of the ROR-P PRS PRS are shown for (A) UK Biobank, (B) the Pathways Study. The mean (standard deviation) of the ROR-P PRS was -27.5 (5.0) in the UK Biobank and -30.9 (5.0) in the Pathways Study.

A.

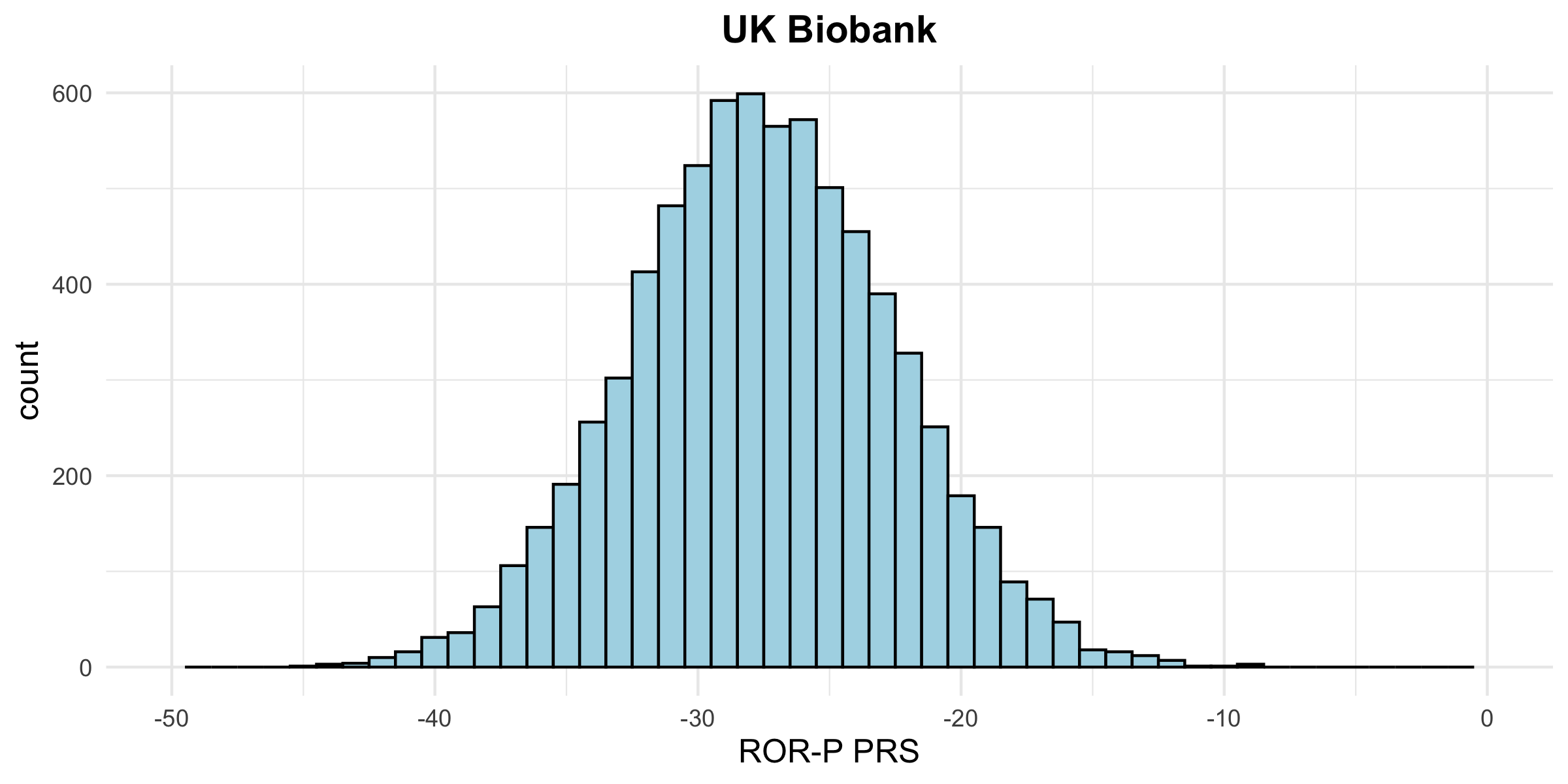

B.

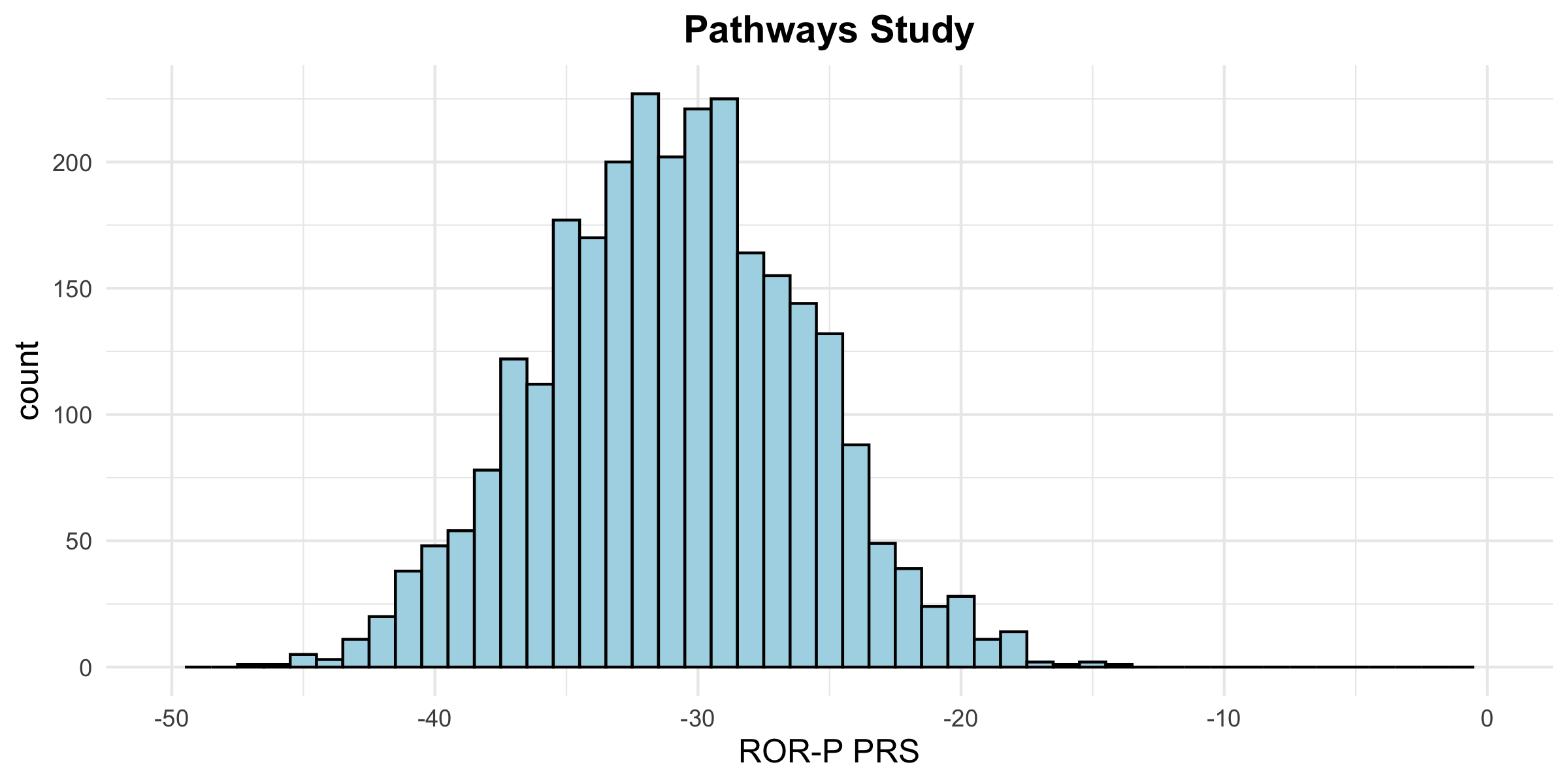

**Figure S4. Calibration of the polygenic risk score for the risk of recurrence score weighted on proliferation (ROR-P PRS) in the UK Biobank**

Predicted versus observed 5-year survival based on the ROR-P PRS are plotted for incident cases in the UK Biobank. Estimates are shown for 7,427 participants divided into 10 strata of predicted survival probability. Black dots correspond to the Kaplan-Meier survival estimate (with 95% error bars) for each stratum of predicted survival. Blue X’s correspond to bias-corrected estimates. The gray line corresponds to ideal calibration. The distribution of predicted survival probabilities is shown along the upper horizontal axis.

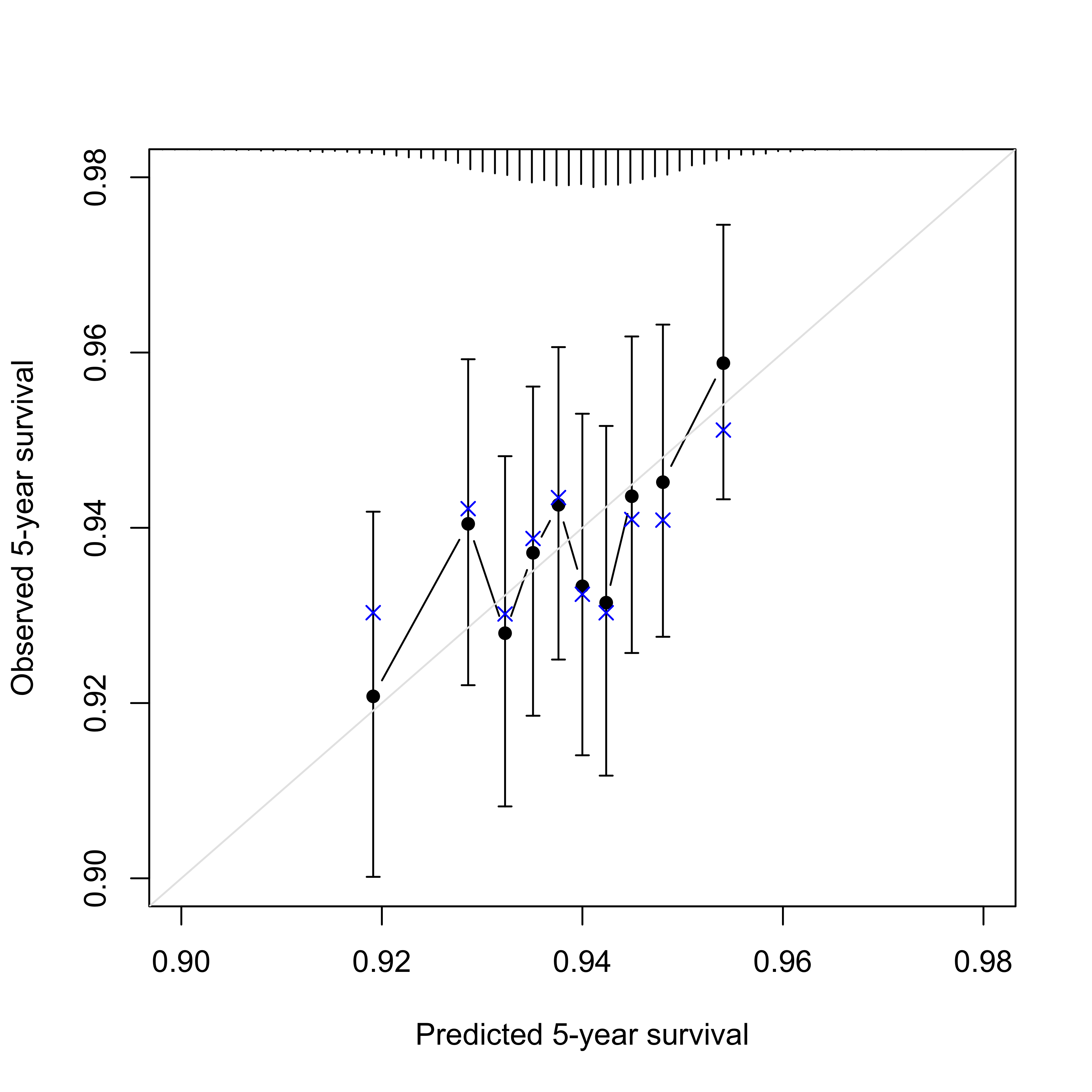

**Figure S5.** **Validation of the polygenic risk score for estrogen-negative versus positive breast cancer (PRS_ER-/ER+_)**

Boxplot of PRS_ER-/ER+_ by immunohistochemical estrogen receptor status in Pathways.

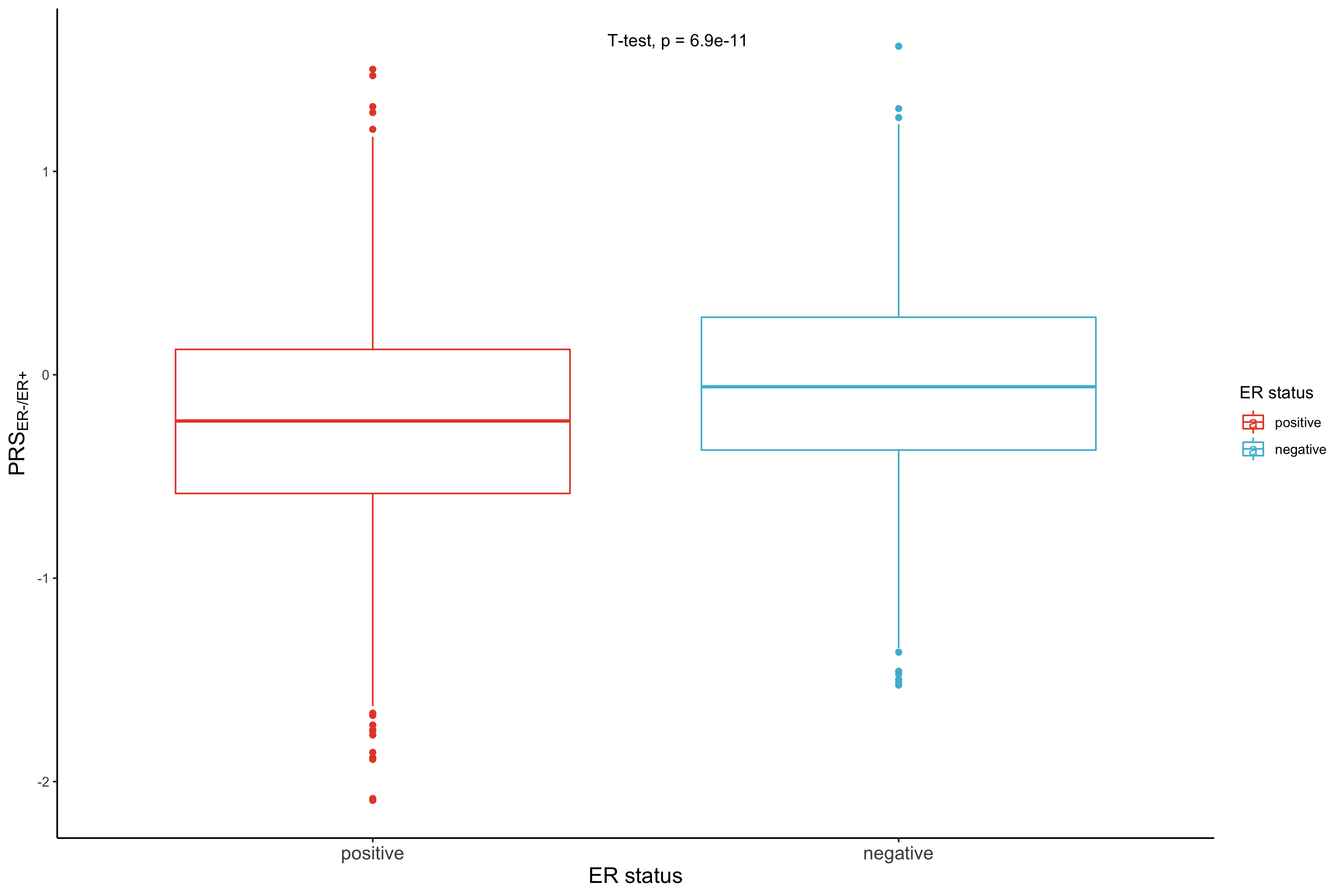

**Figure S6. Correlation between polygenic risk scores for the risk of recurrence score weighted on proliferation (ROR-P PRS) and risk of estrogen-negative versus positive breast cancer (PRS_ER-/ER+_) in the UK Biobank and Pathways Study**

Scatter plot of the normalized ROR-P PRS and log-transformed PRS_ER-/ER+_ in the (A) UK Biobank and (B) the Pathways Study. The Pearson correlation coefficient and p-value are shown.

A.

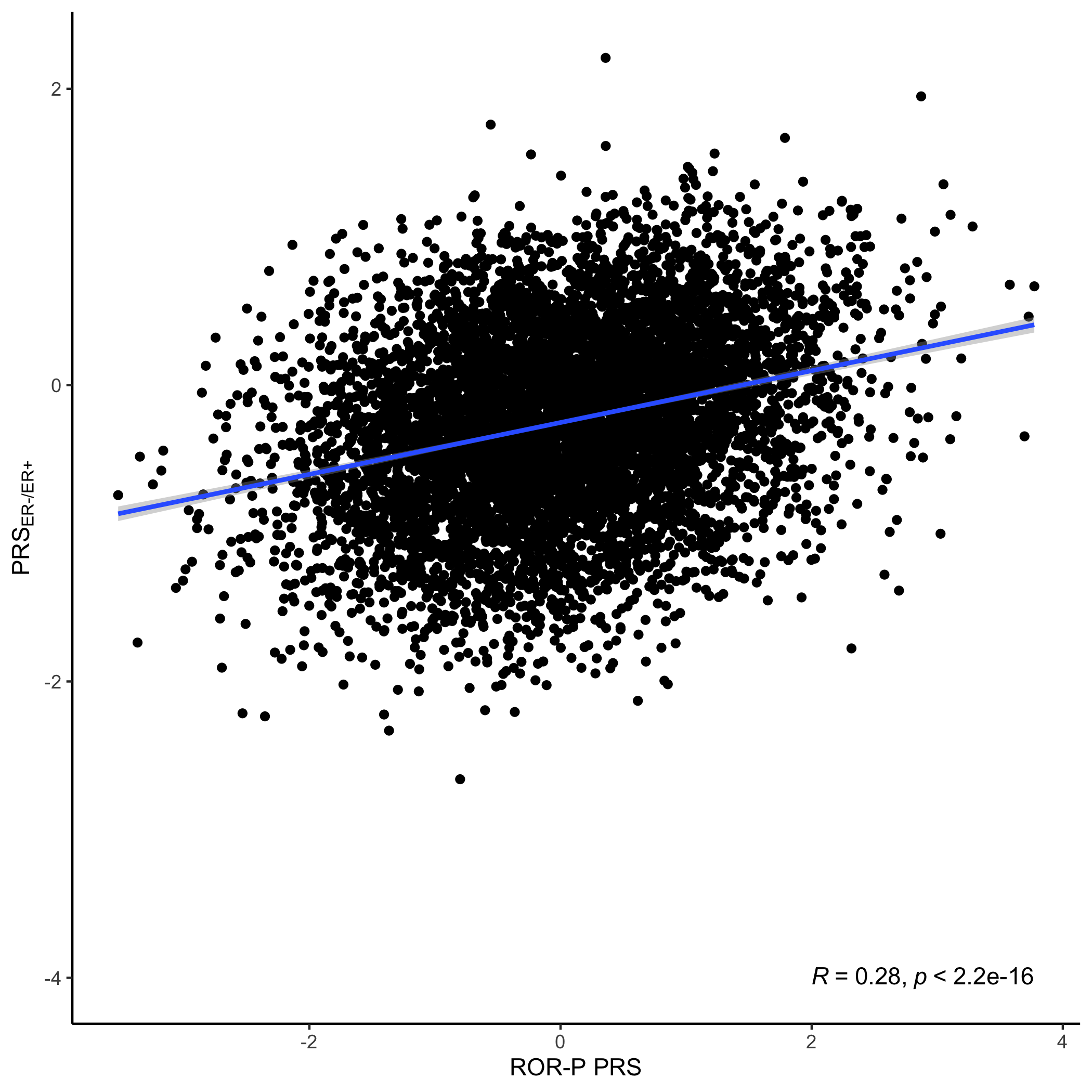

B.

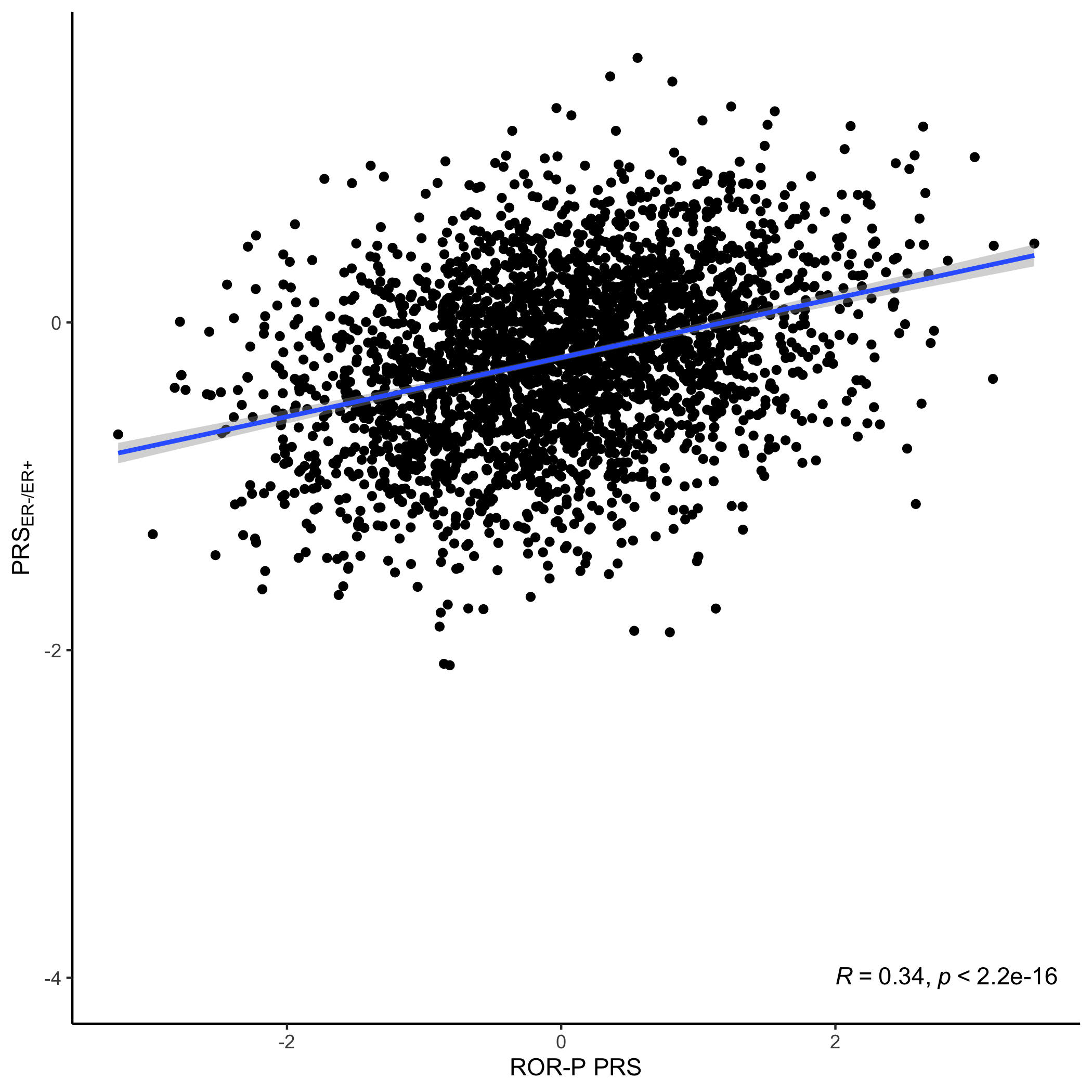

**Figure S7. Performance of the** **polygenic risk score for the risk of recurrence score weighted on proliferation (ROR-P PRS) by estrogen receptor status in the Pathways Study**

(A) Kernel density plot of the ROR-P PRS stratified by estrogen receptor (ER) status. The ROR-P PRS was higher in ER-negative cancers than in ER-positive cancers, mean (standard deviation) of -30.4 (4.9) versus -31.0 (5.0) (t-statistic = -2.2, p = 0.03). (B) Forest plot of the associations between the ROR-P PRS and breast cancer-specific survival in ER-positive cancers (2,386 cases with 177 events) and ER-negative cancers (382 cases with 64 events). Cox proportional hazards models were adjusted for genetic ancestry (principal components 1-10). Hazard ratios per standard deviation are shown.

A.

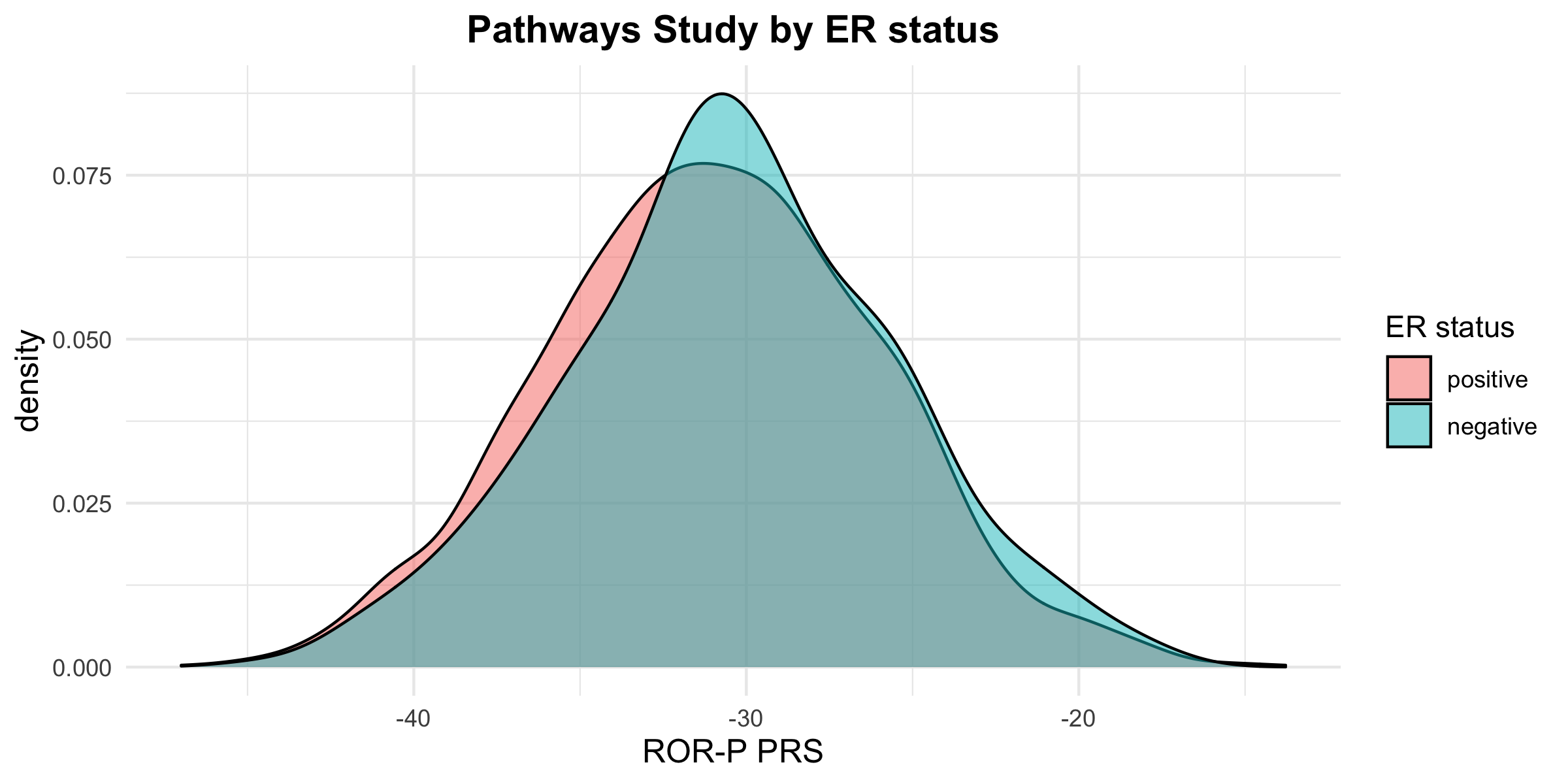

B.

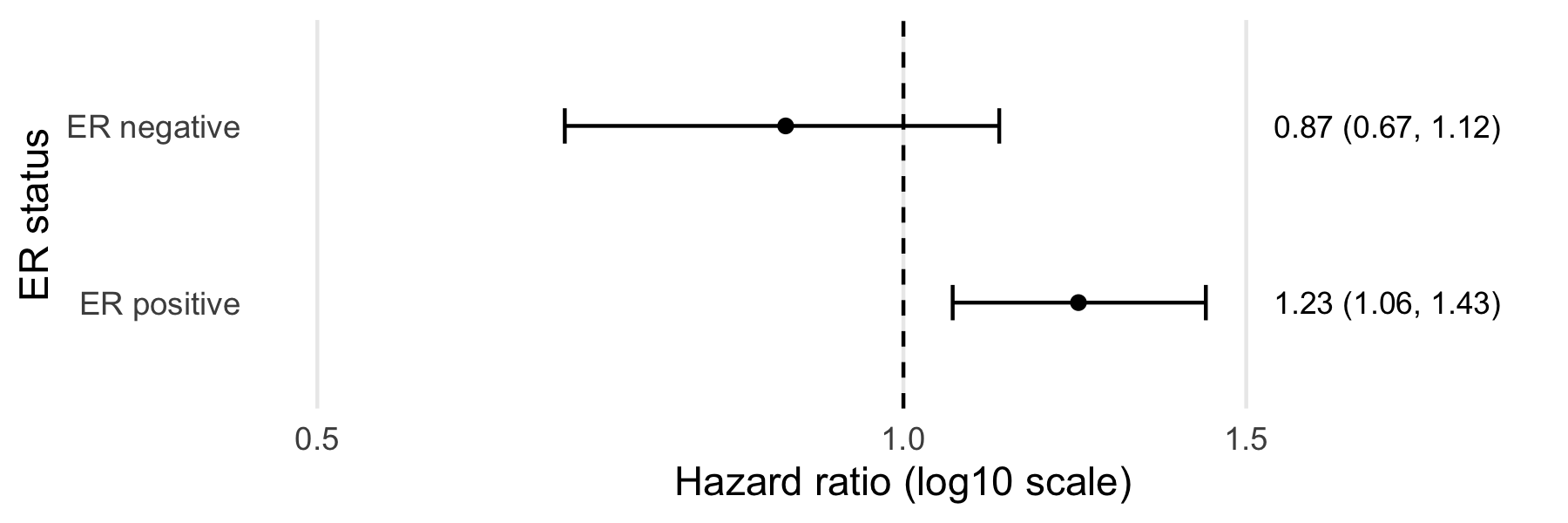
